# Plasma microRNA profiling for malaria disease: association with severity and *P. falciparum* biomass

**DOI:** 10.1101/2020.07.31.20165712

**Authors:** Himanshu Gupta, Mercedes Rubio, Antonio Sitoe, Rosauro Varo, Pau Cisteró, Lola Madrid, Inocencia Cuamba, Alfons Jimenez, Xavier Martiáñez-Vendrell, Diana Barrios, Lorena Pantano, Allison Brimacombe, Mariona Bustamante, Quique Bassat, Alfredo Mayor

**Author notes:** Address for correspondence: Dr. Himanshu Gupta, ISGlobal, Hospital Clínic, Universitat de Barcelona, Carrer Rosselló 153 (CEK Bldg), E-08036 Barcelona, Spain. equal contribution (Himanshu Gupta, Mercedes Rubio). These authors share senior authorship (Quique Bassat, Alfredo Mayor).

## Abstract

Severe malaria (SM) is a major public health problem in malaria-endemic countries. Sequestration of *Plasmodium falciparum* (*Pf*) infected erythrocytes in vital organs and the associated inflammation leads to organ dysfunction. MicroRNAs (miRNAs), which are rapidly released from damaged tissues into the host fluids, constitute a promising biomarker for the prognosis of SM. This study applied next-generation sequencing to evaluate the differential expression of miRNAs in SM compared to uncomplicated malaria (UM). Six miRNAs were associated with *in vitro Pf* cytoadhesion, severity in Mozambican children and *Pf* biomass. Relative expression of hsa-miR-4497 quantified by TaqMan-RT-qPCR, was higher in SM children plasmas compared to that of UM (p<0.048), and again correlated with *Pf* biomass (p=0.033). These findings suggest that different physiopathological processes in SM and UM lead to differential expression of miRNAs and pave the way to future studies aiming to assess the prognostic value of these miRNAs in malaria.

## Introduction

Case fatality rates for *Plasmodium falciparum* (*Pf*) severe malaria (SM) remain unacceptably high in young African children (1). Early detection and prompt treatment of SM are critical to improve the prognosis of sick children. Unfortunately, clinical signs and symptoms in many malaria patients, particularly at the beginning of the infection, may not adequately indicate the potential for the infection to trigger severe or life-threatening disease. Moreover, in malaria-endemic areas, where immunity to malaria is progressively acquired, the detection of peripheral *Pf* parasitemia in sick children does not necessarily prove that malaria is the cause of the severe pathology observed, given that many individuals may carry parasites without expressing clinical malarial disease (2).

Sequestration of *Pf* infected erythrocytes (iEs) (3) in vital organs is believed to constitute a key pathogenic event leading to SM, as has been shown in post-mortem parasite counts in patients who died with cerebral malaria (CM) (4, 5). This extensive sequestration of parasitized erythrocytes in the microvasculature, together with the production of inflammatory mediators, leads to the dysfunction of one or more peripheral organs, such as the lungs (acute respiratory distress syndrome), kidneys (acute kidney injury) or brain (coma) (6, 7). This tissue-specific tropism of *Pf* parasites is mediated by the *Pf* erythrocyte membrane protein-1 (PfEMP1) which can bind to different host receptors on the capillary endothelium, uninfected erythrocytes and platelets (8, 9) such as endothelial receptor of protein C (ePCR), gC1qR, Intercellular adhesion molecule-1, CD36, chondroitin sulfate A or complement receptor 1 (10).

Efforts have been made to identify biomarkers of SM which could be used for prevention of the severity of disease, and for early diagnosis (11). Several biomarkers related to endothelial activation and immune dysfunction have been associated with different malaria-derived severe pathologies (11-14). Plasma levels of histidine-rich protein 2 (HRP2), a parasite-specific protein secreted by the parasite during its blood cycle, has been used as a biomarker of total parasite biomass (circulating and sequestered parasites) (15, 16) and therefore as a prognostic marker of the total parasite biomass and as a better proxy marker for SM than peripheral parasitemia (16). Organ damage and pathological disease states have also been associated with the rapid release of microRNAs (miRNAs) into the circulation, a class of endogenous small non-coding RNAs (18-24 nucleotides) (17). As secreted miRNAs can be detected in biological fluids such as plasma (18), they are currently being explored as promising non-invasive biomarkers to monitor organ functionality and tissue pathophysiological status. The content of miRNAs in the host is influenced by host-pathogen interactions (19). Sequestration of erythrocytes infected with *P. berghei* in mice brains has been demonstrated to modify the miRNA expression in cells (20). Similarly, sequestration of *P. vivax* gametocytes in the bone marrow has been associated with transcriptional changes of miRNAs involved in erythropoiesis (21). The aforementioned evidence suggests that *Plasmodium* parasites, although not able to produce miRNAs (22), could affect the production of organ-specific host miRNAs, pointing towards the potential of these small molecules to detect SM associated organ injury (23) and to confirm the contribution of malaria in the chain of events leading to death through the analysis of postmortem tissues (23).

This study was conducted under the hypothesis that miRNA levels in plasma are differentially expressed among children with severe and uncomplicated malaria due to the parasite sequestration in vital organs of severely ill children. To identify promising biomarkers for SM, a small RNA next-generation sequencing (NGS) was applied to select miRNAs that were differentially expressed by human brain endothelial (HBE) cells exposed to *Pf* iEs selected for cytoadhesion to endothelial receptor of protein C, the main host receptor associated with SM (9), compared to those exposed to non-cytoadherent iEs, and non-infected erythrocytes (niEs), as well as by Mozambican children with SM compared to children with uncomplicated malaria (UM) (Figure 1). miRNAs that were differentially expressed in both analyses, together with the *Pf* biomass-associated miRNAs (correlation coefficient >0.50 (24)), were quantitatively confirmed in an independent validation cohort set of Mozambican children with SM and UM using TaqMan reverse transcriptase quantitative PCRs (RT-qPCRs).

**Figure 1:**
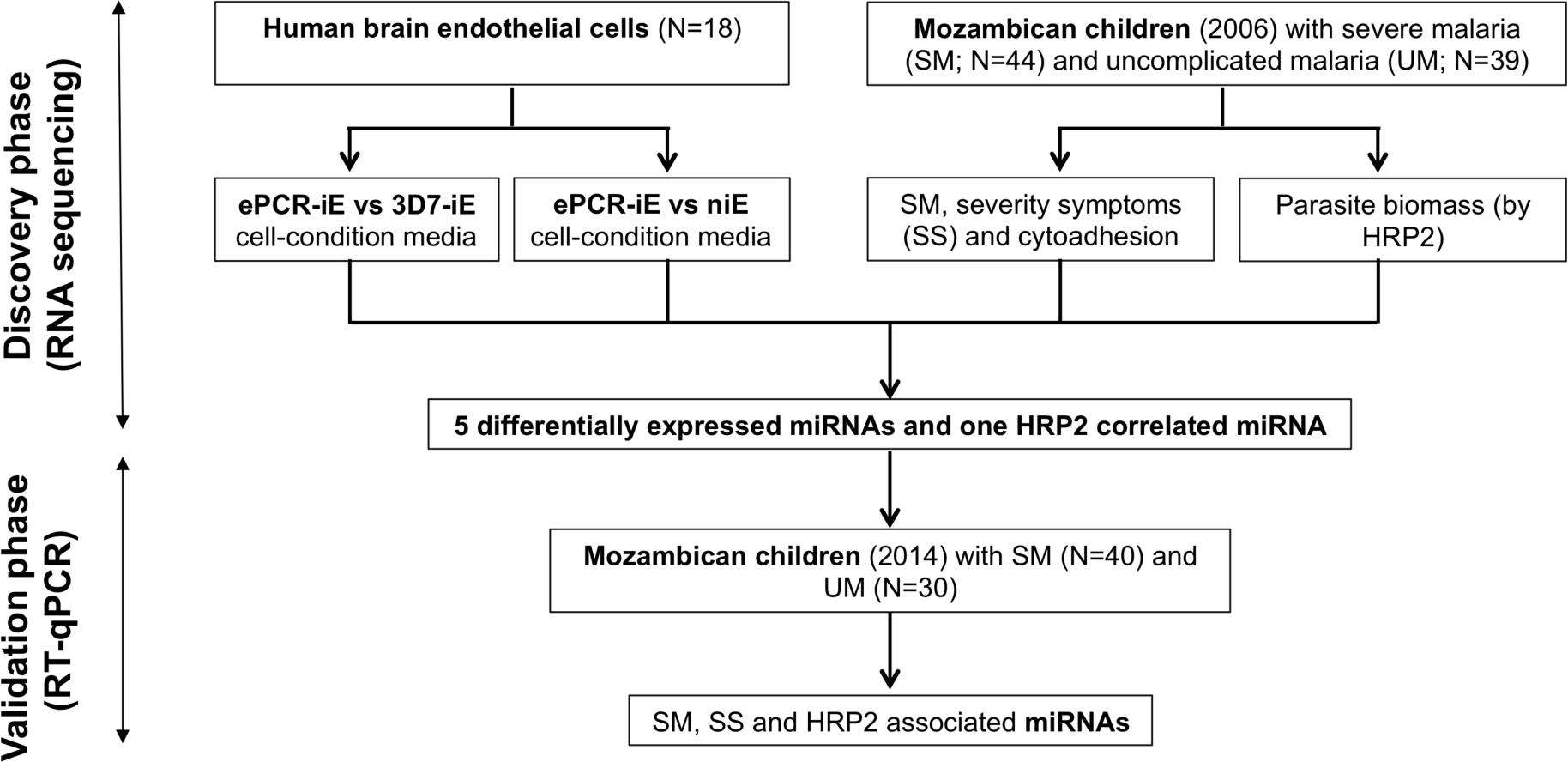
Schematic representation of the study design to identify miRNA based biomarkers of severe malaria. [ePCR: endothelial protein-C receptor (a binding *P. falciparum* strain-FCR3); 3D7: a non-binding *P. falciparum* strain; iE: infected erythrocyte; niE: non-infected erythrocyte; HRP2: Histidine-rich protein 2].

## Material and Methods

### Study population

Plasma samples used to assess miRNA levels were collected in two case-control studies conducted in Manhiça District (southern Mozambique) during 2006 (N=113) and 2014 (N=91). Briefly, the cases were children under five years of age admitted to the Manhiça District Hospital with SM and controls were outpatient children with UM. Details on SM and UM criteria, case management, sample collection and storage, and laboratory blood analysis are included in the Technical Appendix.

The study protocols for each of the two case-control studies from which this analysis was derived were approved by the National Mozambican Ethical Review Committee (Mozambique) and Hospital Clínic (Barcelona, Spain). A signed written informed consent was obtained from all participants’ guardian or parent during the original studies.

### Parasitological determinations

Thick and thin blood films were prepared to quantify *Pf* parasitaemia. Approximately half of a 60µL dried blood drop on Whatman-903 filter paper was used to extract parasite DNA followed by a real-time quantitative PCR (qPCR) targeting the *Pf* 18S rRNA gene (25, 26). HRP2 levels were quantified using commercially available enzyme-linked immunosorbent assay (ELISA) kits and an in-house highly sensitive quantitative bead suspension array (qSA) based on Luminex technology (Technical Appendix).

### *Pf* cytoadhesion assays

Cytoadhesion assays were performed to discover the differential expression of miRNAs. HBE cells were incubated with *Pf*-iEs at the trophozoite stage of the ePCR-binding FCR3 strain (ePCR-iE; which expresses the PfEPM1 protein that binds to ePCR receptor) and the 3D7 strain (3D7-iE; a strain without the protein that binds to ePCR receptor). Non-infected erythrocytes were used as negative control. Details on cytoadhesion assays are included in the Technical Appendix. The cell-conditioned media of each group were collected after 1hr (t1) and 24hrs of stimulation (t24) and subjected to RNA extraction followed by small-RNA sequencing.

### Molecular procedures, gene target prediction and data analysis

RNA was extracted from cell-conditioned media (3ml) and plasma samples (1ml) using the miRNeasy tissues/cells kit and miRNeasy Plasma/Serum kit, respectively, with the use of 5µg UltraPure™ glycogen/sample. Given that the plasma samples were conserved in heparin, RNA was precipitated with LiCl as described elsewhere (27). Purified RNA was subjected to library preparation, pooling and sequencing using a HiSeq 2000 (Illumina) platform following the protocol for small RNAs (28), for more details see the Technical Appendix. A previously published pipeline (28) was used to assess the sequencing quality, identification and quantification of small RNAs, normalization and other species RNA contamination (Technical Appendix). To detect miRNAs and isomiRs, reads were mapped to the precursors and annotated to miRNAs or isomiRs using miRBase version 21 with the miraligner (29). DESeq2 R package v.1.10.1 (R3.3.2) (30) was used to perform an internal normalization.

Fifty µl of plasma from the Mozambican children (2014) with no haemolysis were used for RNA extraction as described above followed by RT-qPCR. Details on RT-qPCR and endogenous controls (ECs) are included in the Technical Appendix. miRNA relative expression levels (RELs) were calculated with the 2^−ΔCt^ method, where ΔCt = [Ct (miRNA) – Mean Ct (ECs)], considering efficiencies of 100% for all the miRNAs and ECs (31).

The selected miRNAs were screened through different gene target prediction programs such as DIANA-microT-CDS, MiRDIP, MirGate, and TargetScan (Technical Appendix).

Differential expression of miRNAs and isomiRs was assessed using DESEq2 and IsomiRs packages in R (29, 32), for more details see the Technical Appendix. All statistical analyses were performed using R3.3.2 and graphs were prepared with GraphPad (Technical Appendix).

## Results

### Discovery phase

#### miRNA expression by HBE cells

The ePCR binding *Pf* strain (FCR3; ePCR-iE) showed higher levels of cytoadhesion to HBE cells (mean of 32.60 iE per 500cells, standard deviation [SD]:4.87) than a non-binding *Pf* (3D7; 3D7-iE) strain (3.20, SD:1.06; p=0.001) and non-infected erythrocytes (3.12, SD:0.39; p=0.001) (Appendix Figure 1). Three replicates of the media collected from each cytoadhesion assay after one (t1) and 24 hours (t24) were sequenced, giving a total of more than 200 million reads per lane, with a mean of 12.10 million reads (SD=13.31) per sample (Table 1; Figure 2A; Appendix Table 1). The mean percentage of miRNAs in the media samples analysed was 4.01% (SD=2.93) and a mean of 203 (SD=93.82) distinct miRNAs (a minimum of 101 and a maximum of 465) were detected (Appendix Table 1). The ten most expressed miRNAs for all samples at t1 and t24 time points are described in Figure 2B. No contamination with RNA from other species was observed.

**Table 1:**
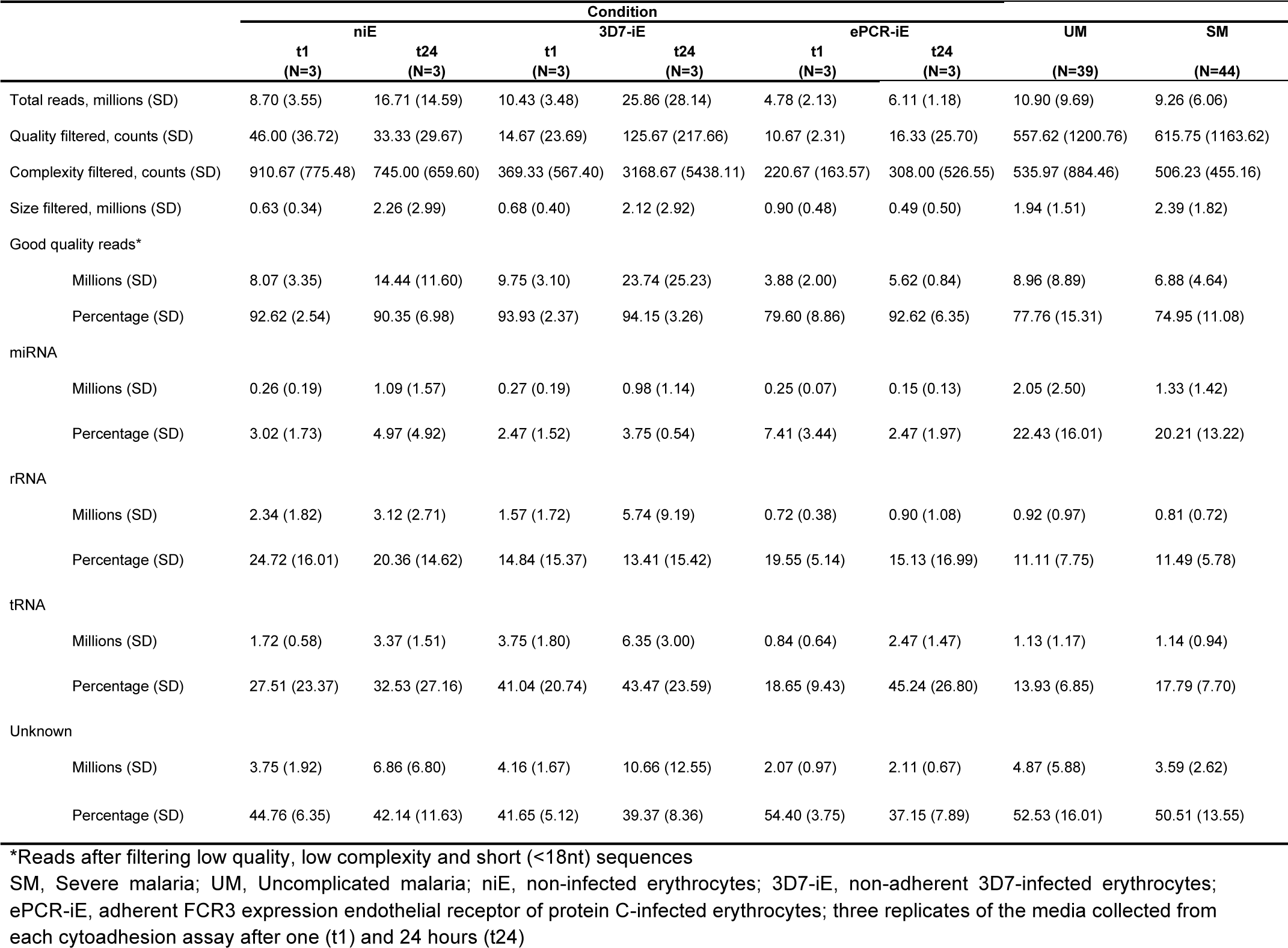
Quality control and mapped reads (mean and standard deviations) in different species of small RNAs obtained from cell-conditioned media of human brain endothelial cells exposed to cytoadherent *P. falciparum* infected and non-infected erythrocytes, and plasma of Mozambican children with severe and uncomplicated malaria.

**Figure 2:**
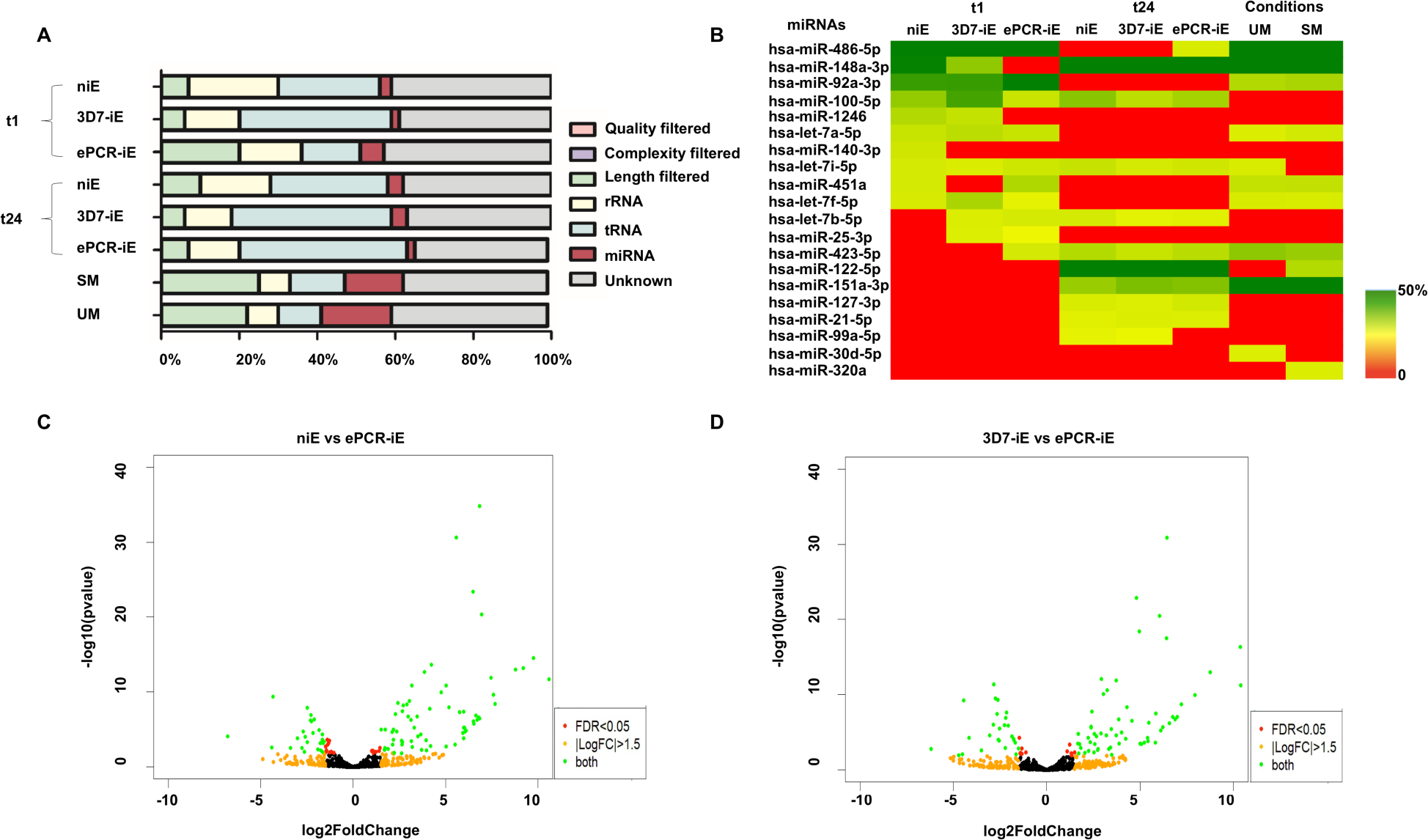
RNA sequencing of human brain endothelial (HBE) cell media and plasma from Mozambican children recruited in 2006. **A**) Percentage of mapped reads in different species of small RNAs, for both *in vitro* and *ex vivo* approaches. **B**) Ten most expressed miRNAs in HBE cell medias and plasmas. Colour coded cells show the percentage of each assay/condition (columns) for each miRNA (rows). Volcano plot of differentially expressed miRNAs in **C**) cell-condition media of non-infected erythrocytes (niE) versus cell-condition media of infected erythrocytes with the FCR3-ePCR strain (ePCR-iE) incubated with HBE cells and **D**) cell-condition media of infected erythrocytes with 3D7 strain (3D7-iE) versus cell-condition media of infected erythrocytes with the FCR3-ePCR strain (ePCR- iE) incubated with HBE cells. Both comparisons (C and D) were adjusted for multiple testing by the Benjamini-Hochberg method. Negative log2FoldChanges indicates overexpression in ePCR-iE samples. (SM: severe malaria; UM: uncomplicated malaria).

One hour after incubating the HBE cells with *Pf* infected and non-infected erythrocytes, 111 miRNAs were found to be differentially expressed in cell-condition media of niE and ePCR-iE, 76 of them being downregulated and 35 upregulated in ePCR-iE compared to niE (Figure 2C; Appendix Table 2). At this same time point, 100 miRNAs were differentially expressed in cell-condition media of 3D7-iE and ePCR-iE, 67 were downregulated and 33 upregulated in ePCR-iE compared to 3D7-iE (Figure 2D; Appendix Table 3). Overall, 89 miRNAs were differentially expressed in ePCR-iE compared to both niE and 3D7-iE, 28 and 61 of which were upregulated and downregulated, respectively, in ePCR-iE. There were no differentially expressed miRNAs between niE and 3D7-iE cell-condition media. At t24, only hsa-miR-451a was significantly upregulated in cell-condition media of ePCR-iE with respect to niE and 3D7-iE. There were no significantly different miRNAs found between niE and 3D7-iE cell-condition media. All differentially expressed isomiRs were from the selected miRNAs and none of them presented any modifications in the seed region.

**Table 2:** Characteristics of Mozambican children with severe and uncomplicated malaria recruited in 2006 (discovery) and 2014 (validation).

|  |  | 2006 |  |  | 2014 |  |  |
| --- | --- | --- | --- | --- | --- | --- | --- |
|  |  | UM<br>N=39 | SM<br>N=44 | p value | UM<br>N=30 | SM<br>N=40 | p value |
| Age<sup>#</sup> (year), mean (SD) |  | 2.3 (1.1) | 2.4 (1.3) | 0.671 | 2.2 (1.3) | 2.8 (1.2) | 0.419 |
| Sex, n (%) |  |  |  |  |  |  |  |
|  | Men | 24 (62) | 28 (64) | 1.000 | 18 (60) | 21 (52.5) | 0.532 |
|  | Women | 15 (38) | 16 (36) |  | 12 (40) | 19 (47.5) |  |
| HRP2 (ng/ml), GM (SD) |  | 71.3 (10.7) | 331.4 (40.7) | <0.001 | 24.1 (4.9) | 78.7 (12.2) | 0.038 |
| qPCR (parasites/μl), GM (SD) |  | 2084.9<br>(302.5) | 7976.1<br>(1079.6) | 0.004 | 72845.9<br>(7193.9) | 94099.6<br>(8716.0) | 0.549 |
| Splenomegaly, n (%) |  |  |  |  |  |  |  |
|  | No | 33 (85) | 21 (48) | 0.001 | ND | 27 (67.5) | NA |
|  | Yes | 6 (15) | 23 (52) |  | ND | 13 (32.5) |  |
| Hepatomegaly, n (%) |  |  |  |  |  |  |  |
|  | No | 38 (97) | 35 (80) | 0.016 | ND | 35 (87.5) | NA |
|  | Yes | 1 (3) | 9 (20) |  | ND | 5 (12.5) |  |
| Hyperlactatemia, n (%) |  |  |  |  |  |  |  |
|  | No | 10 (26) | 5 (11) | 0.152 | 26 (86.7) | 27 (67.5) | 0.064 |
|  | Yes | 29 (74) | 39 (89) |  | 4 (13.3) | 13 (32.5) |  |
| Temperature (°C), mean (SD) |  | 38.0 (1.6) | 38.5 (1.1) | 0.093 | 38.0 (1.3) | 38.2 (1.4) | 0.437 |
| Weight (Kg), mean (SD) |  | 11.3 (2.8) | 11.0 (2.8) | 0.599 | 12.3 (2.9) | 12.7 (3.3) | 0.476 |
| Platelets (10<sup>9</sup>/L), mean (SD) |  | 156.7 (86.8) | 115.8 (66.8) | 0.018 | 149.0 (89.7) | 95.3 (69.3) | 0.001 |
| Glucose<sup>a</sup> (mM), mean (SD) |  | 6.2 (1.5) | 5.9 (1.8) | 0.391 | 6.6 (1.3) | 6.0 (2.6) | 0.165 |
| WBC (10<sup>9</sup>/L), mean (SD) |  | 9.9 (4.1) | 10.2 (3.9) | 0.774 | 9.7 (3.8) | 9.6 (5.0) | 0.929 |
| Neutrophils<sup>c</sup> (%), mean (SD) |  | 54.1 (16.7) | 54.4 (14.3) | 0.940 | 50.7 (20.6) | 58.9 (13.7) | 0.447 |
| Lymphocytes<sup>d</sup> (%), mean (SD) |  | 39.4 (17.9) | 36.3 (12.6) | 0.374 | 26.1 (17.1) | 25.6 (12.2) | 0.995 |
| Lactate (mM), mean (SD) |  | 3.0 (1.7) | 4.7 (3.6) | 0.009 | 2.8 (2.2) | 3.6 (2.4) | 0.035 |
| Severe malaria syndromes, n (%) |  |  |  |  |  |  |  |
|  | Prostration |  | 33 (75.0) |  |  | 30 (75.0) |  |
|  | Acute respiratory distress |  | 18 (40.9) |  |  | 19 (47.5) |  |
|  | Severe anaemia |  | 17 (38.6) |  |  | 7 (17.5) |  |
|  | Multiple seizures |  | 11 (25.0) |  |  | 24 (60.0) |  |
|  | Cerebral malaria |  | 2 (4.5) |  |  | 7 (17.5) |  |
|  | Hypoglycaemia |  | 2 (4.5) |  |  | 2 (5.0) |  |

Continuous data = Mann Whitney test; categorical data = Chi-square test
SM, Severe malaria; UM, Uncomplicated malaria; HRP2, Histidine-rich protein 2; WBC, White blood cells; GM, Geometric mean; SD, Standard deviation; ND, not determined; NA, not applicable
<sup>#</sup> No data of 1 sample (UM=1) in 2014 study
<sup>a</sup> No data of 3 sample (SM=2; UM=1) in 2014 study
<sup>c</sup> No data of 4 samples (SM=4) in 2014 study
<sup>d</sup> No data of 3 samples (SM=3) in 2014 study

**Table 3:** Association of miRNA levels with severe malaria, symptoms of severity and *Plasmodium falciparum* cytoadhesion. Positive FoldChange indicates overexpression in severe malaria and symptoms of severity compared to uncomplicated malaria as well as parasites showing cytoadhesion compared to none.

| miRNA | baseMean | log2FoldChange | padj |
| --- | --- | --- | --- |
| **Clinical data** |  |  |  |
| **Severe malaria (N=44 vs 39)** |  |  |  |
| hsa-miR-122-5p | 19929.69 | 1.67 | 0.001 |
| **Severe anaemia (N=17 vs 39)** |  |  |  |
| hsa-miR-4492 | 17.34 | 2.81 | 0.046 |
| hsa-miR-4497 | 293.66 | 2.18 | 0.046 |
| **Prostration (N=33 vs 39)** |  |  |  |
| hsa-miR-122-5p | 20677 | 1.89 | 0.001 |
| hsa-miR-6087 | 5.36 | 2.39 | 0.033 |
| hsa-miR-511-5p | 126.67 | 1.36 | 0.040 |
| **Acidosis or respiratory distress (N=18 vs 39)** |  |  |  |
| hsa-miR-122-5p | 13367.43 | 2.21 | <0.001 |
| hsa-miR-4497 | 272.39 | 2.05 | 0.07 |
| **Cytoadhesion data** |  |  |  |
| **Platelet-mediated agglutination (N=50 vs 19)** |  |  |  |
| hsa-miR-3158-3p | 1180.96 | -2.26 | <0.001 |
| hsa-miR-320a | 22005.69 | -1.48 | 0.001 |
| hsa-miR-4492 | 18.33 | 2.78 | 0.002 |
| hsa-miR-1290 | 1011.34 | -1.38 | 0.014 |
| hsa-miR-320b | 1191.44 | -1.23 | 0.014 |
| hsa-miR-320c | 408.32 | -1.29 | 0.014 |
| hsa-miR-1246 | 3907.45 | -1.32 | 0.019 |
| hsa-miR-6741-5p | 48.11 | -1.81 | 0.023 |
| hsa-miR-1228-5p | 82.73 | -1.88 | 0.023 |
| hsa-miR-3195 | 16.35 | 2.21 | 0.023 |
| hsa-miR-7706 | 334.86 | -1.00 | 0.023 |
| **gC1qR (N=35 vs 34)** |  |  |  |
| hsa-miR-1-3p | 622.35 | 2.09 | 0.003 |

Total number of miRNAs were 1450 in RNA sequencing data; baseMean = Mean normalized expression of the miRNAs in all the samples; padj = adjusted for multiple testing by the Benjamini-Hochberg method

#### miRNAs expression in plasmas from Mozambican children with malaria of varying severity

Out of 113 plasma samples collected from Mozambican children with SM (N=57) and UM (N=56) in 2006, 11 samples were discarded because of haemolysis (N=5; OD_414_>0.2) (33), and because no peak was observed between 133-150 nucleotides (typical size for miRNAs plus library adaptors) on the bioanalyzer results after library preparation (N=6). Among the 102 sequenced samples (SM=53; UM=49), 19 samples (9 SM; 10 UM) were further excluded because of the low number of miRNA reads (<10,000 reads). Finally, samples from 83 children (44 with SM and 39 with UM) were included in the analysis (Table 2). The characteristics of Mozambican children are presented in Table 2.

The sequencing of the 83 plasma samples yielded a mean of 9.42 million reads (SD=6.4) per sample (Figure 2A; Table 1; and Appendix Table 4). The mean percentage of miRNAs per plasma samples was 20.5% (SD=13.2), with a mean of 395 (SD=169) distinct miRNAs detected (a minimum of 116 and a maximum of 786; Appendix Table 4). The total number of miRNAs detected across samples was 1450. The ten most expressed miRNAs can be found in Figure 2B. No contamination with RNA from other species was observed.

hsa-miR-122-5p was found upregulated in children with SM (Table 3). In the sub-analysis by signs of severity, five miRNAs were found associated with SA, prostration and ARD (Table 3). Twelve miRNAs were found associated with PM-agglutination and cytoadhesion to g1CqR (Table 3). No associations were observed between miRNA counts and rosetting, CD36 and CD54. After adjusting for multiple comparisons, three (hsa-miR-10b-5p, hsa-miR-378a-3p and hsa-miR-4497) out of the 1450 miRNAs (identified in RNA sequencing data) were found to correlate with HRP2 levels determined by qSA (Spearman analysis; Figure 3). Similar correlations were observed when HRP2 levels were determined by ELISA (Appendix Table 5). miRNAs were neither associated with hepatomegaly nor with splenomegaly. All differentially expressed isomiRs between children with SM and UM belong to the differentially expressed miRNAs, with no modifications in the seed region.

**Figure 3:**
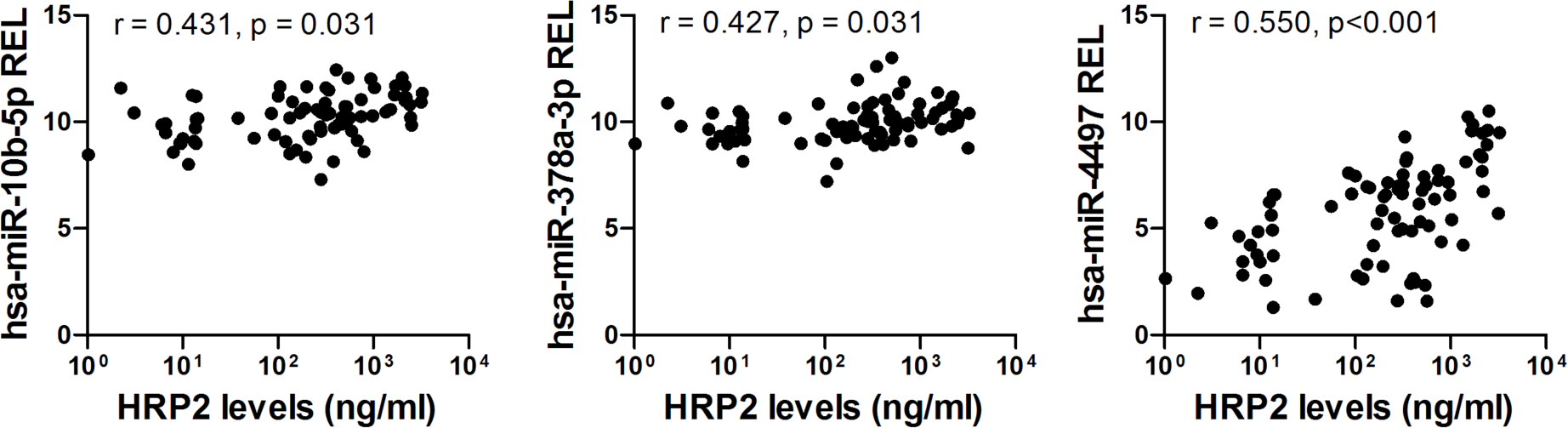
Spearman correlations between HRP2 levels and miRNA relative expression levels (RELs) in plasma samples from Mozambican children recruited in 2006. HRP2 levels and miRNA RELs were log transformed. The correlation analysis was adjusted for multiple testing by the Benjamini-Hochberg method.

#### Validation cohort

Among the 89 miRNAs differentially expressed in cell-condition media of HBE cells exposed to niE and 3D7-iE compared to ePCR-iE, 5 miRNAs were further confirmed to be differentially expressed between children with SM and UM. These five miRNAs (hsa-miR-122-5p, hsa-miR- 320a, hsa-miR-1246, hsa-miR-1290 and hsa-miR-3158-3p) along with hsa-miR-4497 miRNA, which had a correlation coefficient with HRP2 higher than 0.5 (Figure 3), were selected for TaqMan-RT-qPCRs validation in an independent cohort of children with SM and UM recruited in 2014. Among the 91 plasma samples collected from these children, 21 were discarded because of haemolysis (OD_414_>0.2) (33). Out of the 70 remaining samples, 40 and 30 samples were collected from children with SM and UM, respectively (Table 2). The characteristics of Mozambican children are presented in Table 2.

All samples tested by RT-qPCR amplified the exogenous control (ath-miR-159a) with a Ct value<18 and a coefficient of variance (CV) <5%, suggesting the correct RNA extraction and cDNA preparation. hsa-miR-191-5p (CV=4.8%, basemean=3953.3, log_2_fold change (FC)=-0.02 and SD=0.56), hsa-miR-30d-5p (CV=4.9%, basemean=14172.31, FC=0.01 and SD=0.61) and hsa-miR- 148a-3p (CV=5%, basemean=111593.08, FC=0.11 and SD=0.82) were selected as a panel of ECs for RT-qPCR analysis. Among these three, the NormFinder stability value was 0.044 for the combination of hsa-miR-30d-5p and hsa-miR-191-5p, and were thus selected as ECs. No statistically significant differences were found when Ct values of exogenous and two endogenous controls were compared between SM and UM samples (Appendix Figure 2). Standard curves for all miRNAs (ECs and selected miRNAs) were performed, giving efficiencies between 91.1% - 103.8% (Appendix Table 6), which were assumed as 100% to calculate the relative expression values using the 2^−ΔCt^ method (31).

The relative expression levels of hsa-miR-3158-3p and hsa-miR-4497 were significantly higher in children with SM than UM (p<0.05) as shown in Figure 4. hsa-miR-3158-3p levels were higher in children with prostration, multiple seizures and ARD compared to UM (p<0.05; Figure 5). Severe anaemia and ARD symptoms were associated with higher hsa-miR-4497 levels (p<0.05; Figure 5). No such associations were observed for CM and hypoglycaemia. RELs of hsa-miR-3158-3p and hsa-miR-4497 were found positively correlated with HRP2 levels quantified by qSA (p<0.05; Figure 6), with similar correlations observed when HRP2 levels were determined by ELISA (Appendix Table 5).

**Figure 4:**
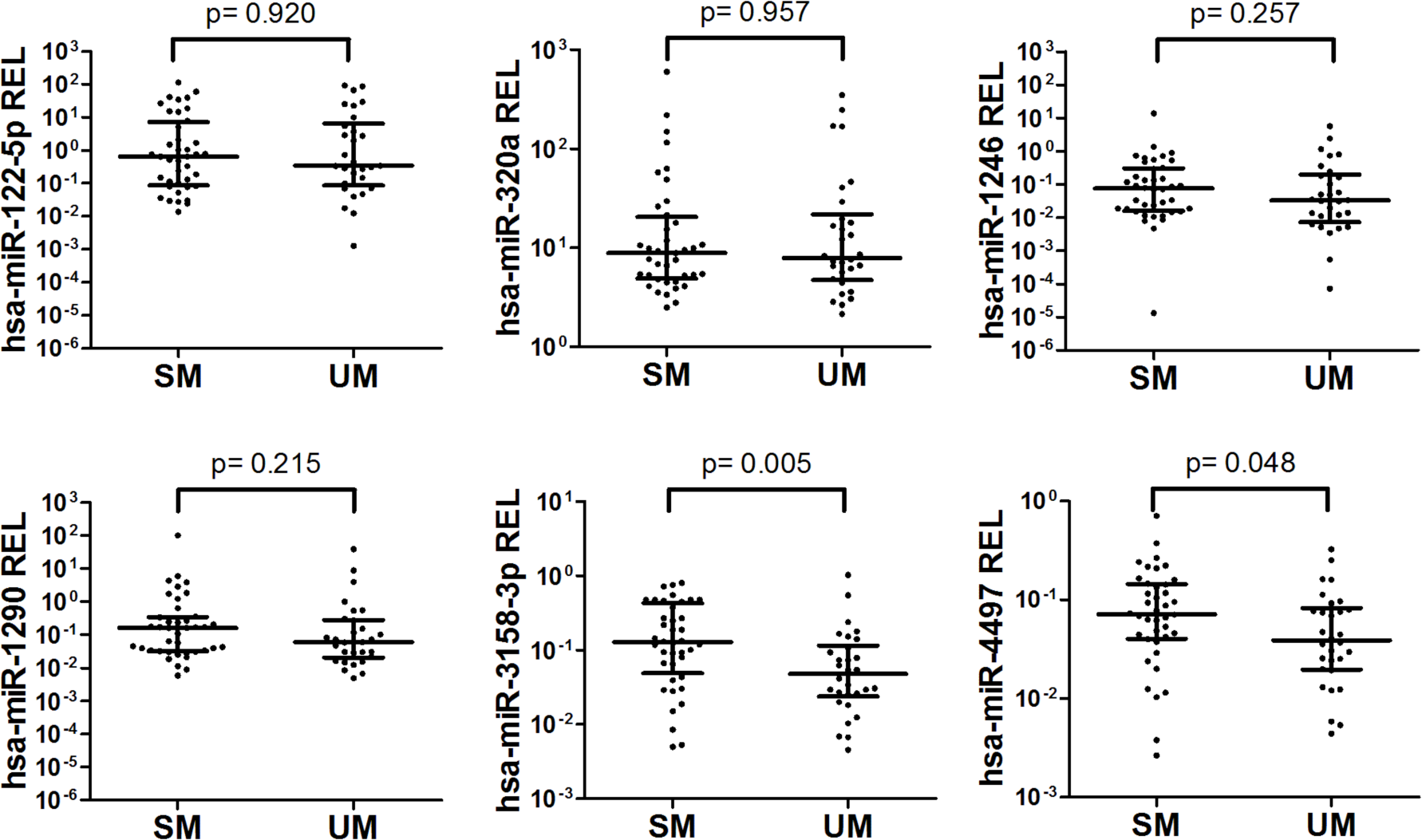
miRNA validation in plasma samples of Mozambican children recruited in 2014. Relative expression levels (RELs) were calculated with respect to the mean of two endogenous controls (hsa-miR-30d-5p and hsa-miR-191-5p) and compared between children with severe malaria (SM) and uncomplicated malaria (UM). Statistical differences were obtained from Mann-Whitney U test. T bars represent median and Interquartile Ranges (IQR).

**Figure 5:**
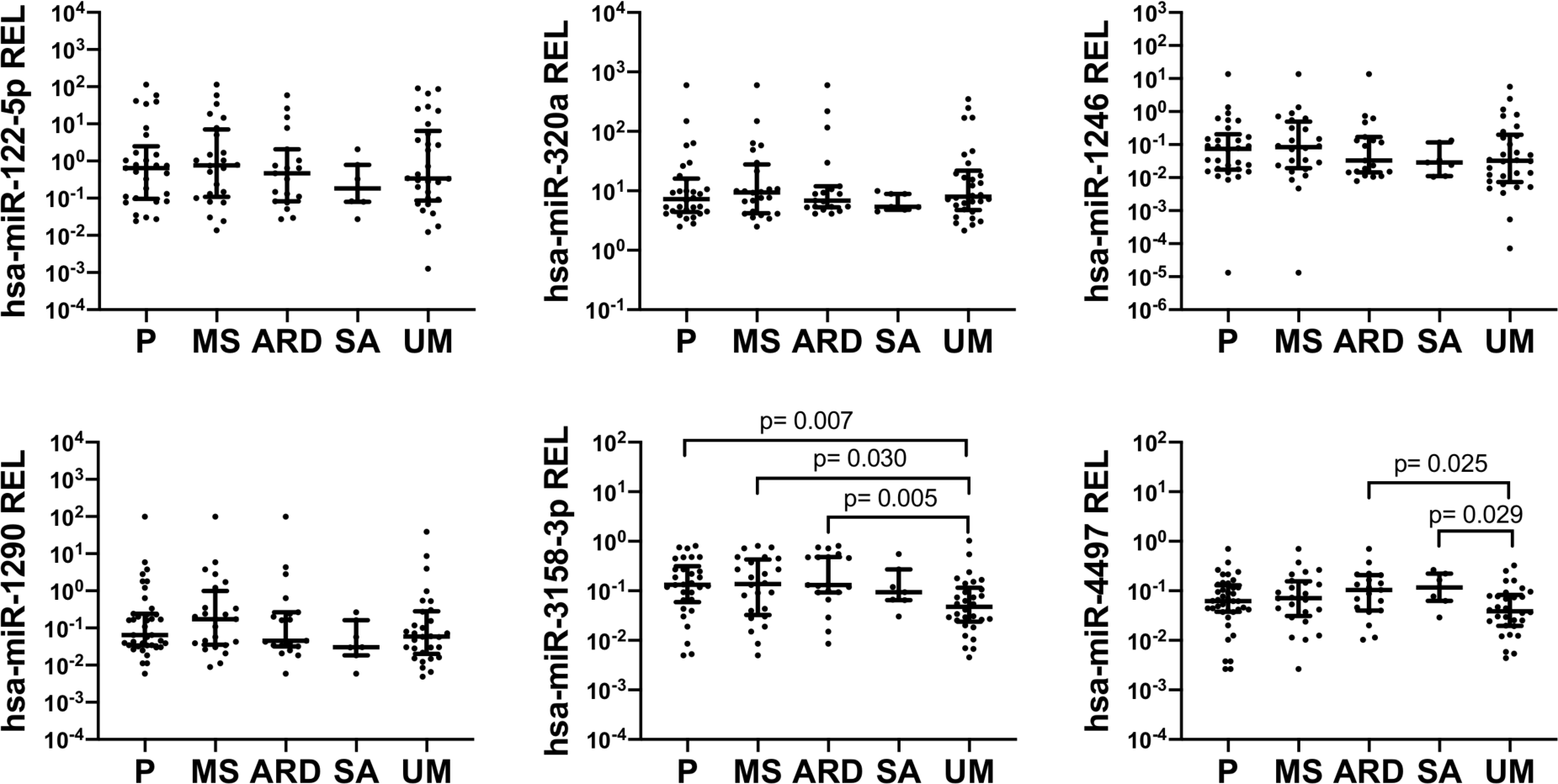
Association of miRNA levels with symptoms of severity. Relative expression levels (RELs) were calculated with respect to the mean of two endogenous controls (hsa-miR-30d-5p and hsa-miR-191-5p) and compared between children with uncomplicated malaria (UM) and symptoms of severity. Distributions were compared using Mann-Whitney U test. T bars represent median and Interquartile Ranges (IQR). P values are shown for significant comparisons. [Prostration (P), Multiple seizures (MS), Acidosis or acute respiratory distress (ARD), Severe anaemia (SA)].

**Figure 6:**
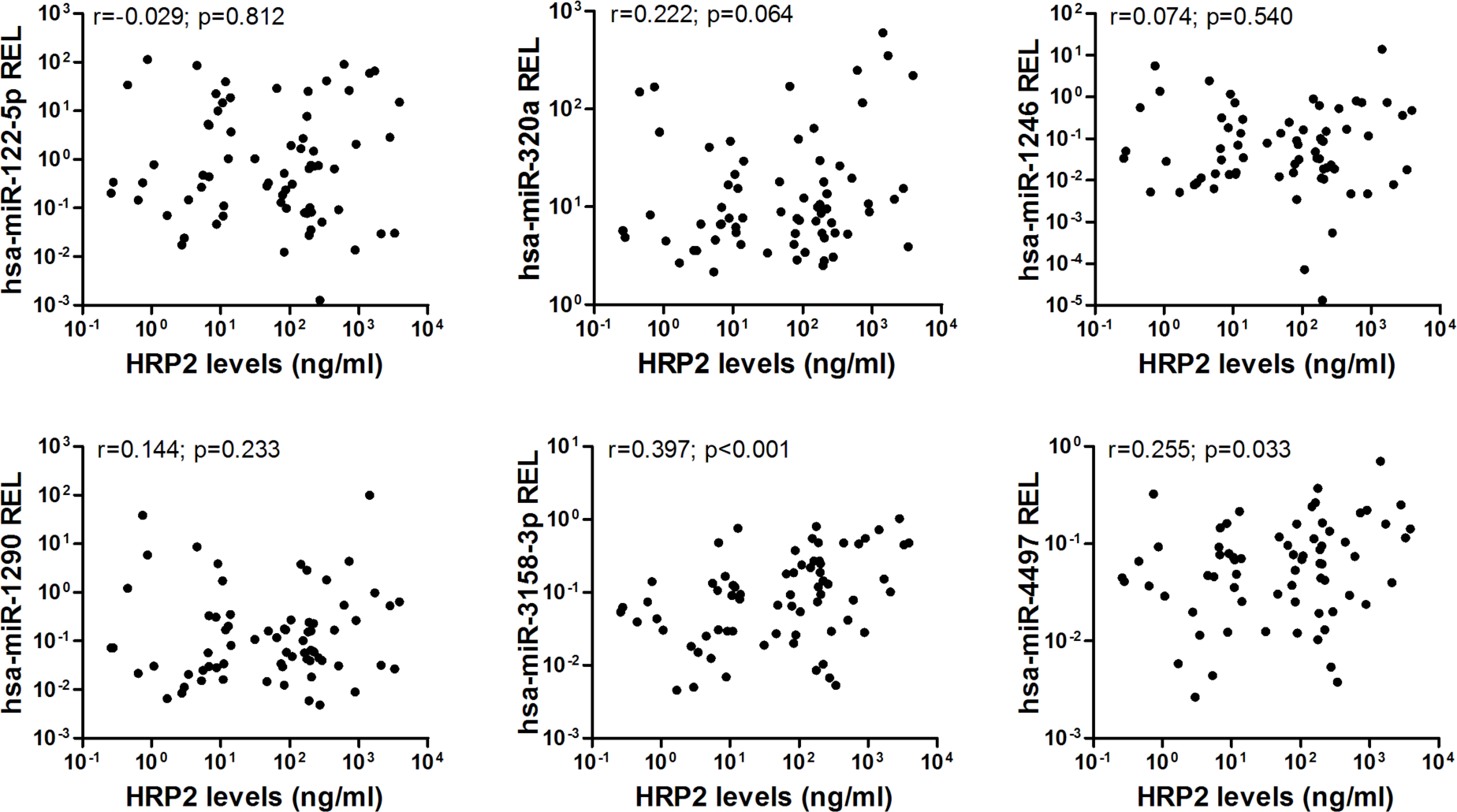
Spearman correlations between HRP2 levels and miRNA relative expression levels (RELs) in plasma samples from Mozambican children recruited in 2014. HRP2 levels and miRNA RELs were log transformed.

#### miRNA gene target prediction

A total of 87 putative targets for hsa-miR-3158-3p and hsa-miR-4497 miRNAs were identified, none of which were shared by both miRNAs (Appendix Table 7). Forty-five experimentally validated mRNA targets were predicted for hsa-miR-3158-3p and 42 for hsa-miR-4497. The predicted targets were found to be involved in a broad range of biological processes (Appendix Table 8). However, significance was lost when adjusted by the Benjamini-Hochberg method. None of the target genes were clustered under the KEGG pathway with p value <0.05.

## Discussion

As microRNAs can reflect disease states and organ damage due to their specificity to cell type (17), they have the potential to provide a new screening method for early detection of pathological *Pf* sequestration and may consequently become an effective prognosis tool for severe malaria. Moreover, the detection of miRNAs associated with organ damage in host biofluids may provide an alternative to post-mortem autopsies for determining the presence of parasites in host vital organs. This approach creates new opportunities to develop malaria diagnostic tools that can guide treatment decisions, and to understand the role of human miRNAs in several disease conditions (23).

In the discovery phase, 89 miRNAs were found to be differentially expressed in the media of HBE cells after incubation with an ePCR-cytoadherent *Pf* strain as compared with non-cytoadherent parasites and non-infected erythrocytes. In addition, fifteen miRNAs in plasma samples obtained from Mozambican children were associated with SM, with specific severity symptoms, and with the cytoadherent *Pf* phenotype, compared to UM and non-cytoadherent parasites. In the validation phase, the higher abundance of hsa-miR-3158-3p and hsa-miR-4497 in SM children compared to children with UM was further confirmed. Prostration, multiple seizures, SA and ARD symptoms of severity were associated with higher levels of hsa-miR-3158-3p and hsa-miR-4497. hsa-miR-4497 levels were also positively correlated with the parasite biomass as quantified by the levels of HRP2 both in the discovery and validation phases. Overall, these findings suggest that different physiopathological processes in SM and UM lead to differential expression of miRNAs in plasma.

HBE cells released a high number of the miRNAs when they were stimulated with an ePCR-binding *Pf* strain within the first hour of incubation. After 24 hours the system stabilizes and only one miRNA (hsa-miR-451a) was found at higher levels in cell-conditioned media of HBE cells incubated with an ePCR-binding strain compared to cells stimulated with non-adherent (3D7-iE) or non-infected erythrocytes. miR-451 has been implicated in translocation to form a chimera with *Plasmodium* mRNAs to block their translation (34), and also found to be abundant in sickle erythrocytes (35). In addition, it was shown that parasites could reduce miR-451 levels in host fluids (36). However, it was not confirmed in plasmas from Mozambican children in this study. Five miRNA levels were higher in children with SM and severity symptoms (prostration, SA and ARD) compared to UM cases. *Pf* cytoadhesion phenotypes (PM-agglutination and cytoadhesion to gC1qR) were also associated with the differential expression of miRNAs, suggesting that the interaction between PfEMP1 and host receptors leads to the secretion to plasma of specific miRNAs. Moreover, three miRNAs (hsa-miR-10b-5p, hsa-miR-378a-3p and hsa-miR-4497) were positively correlated with HRP2 levels.

Six candidate miRNAs that were identified in the discovery phase were selected to determine the validity of the previous results in an independent cohort of Mozambican children. The relative expression of hsa-miR-3158-3p and hsa-miR-4497 was significantly higher in children with SM compared to UM, with hsa-miR-3158-3p levels being higher in children with prostration, multiple seizures as well as ARD, and hsa-miR-4497 in children with SA and ARD. To our knowledge, hsa- miR-3158-3p, which is widely expressed in skin, spleen, kidney and brain tissues (37), has been associated with bipolar disorders (38), but not with other infectious diseases. Further validation is required for hsa-miR-3158-3p, as the levels of this miRNA were found to be downregulated in the plasma from Mozambican children recruited in 2006 with positive PM-agglutination compared to no PM-agglutination, a *Pf* cytoadhesion phenotype which has been associated with malaria severity (39). However, the positive correlation of hsa-miR-4497 with HRP2 levels, which was consistently observed in the cohort of children from 2006 and 2014, suggesting that increasing parasite biomass associated with parasite sequestration may lead to higher levels of secretion of this specific miRNA by damaged tissues. hsa-miR-4497 is widely expressed in the lymph nodes, spleen, kidney and liver tissues (37). Overall, this study shows that hsa-miR-4497, which is also associated with SM, might be an interesting proxy marker of malaria severity. However, hsa-miR-4497 was found as a tumour suppressor (40), and associated with *Mycobacterium tuberculosis* infection (41). Therefore, longitudinal studies are required to assess the prognostic value of this miRNA, as well as to estimate its differential expression in children with severity due to non-malarial infections.

Few of the most expressed miRNAs found in the present study, which represent a 70% of the total miRNA counts in plasma samples, have been reported as highly abundant in plasma samples previously (28, 42). According to public data deposited in the miRmine database (43), hsa-miR- 486-5p and hsa-miR-451a are the two most abundant miRNAs in plasma and were also present in the list of ten most expressed miRNAs of this study. Although there is no data available on miRNAs from cell-conditioned media of HBE cells, miRNA data from other cell types, such as primary tissue explants, primary stromal cells and breast cancer cell lines, also show low miRNA yield (44), similar to this study. This observation indicates that RNA sequencing data obtained in this study is of good quality and can be used for posterior analysis with high confidence. However, this study has several limitations. First, only HBE cells and ePCR-binding parasites were utilized for the *in vitro* assay, and therefore miRNAs produced by other parasite-host interactions contributing to SM may have been missed. Second, plasma samples used in this study were collected retrospectively. Therefore, factors prior to small RNA sequencing and TaqMan-RT-qPCRs, such as time taken between centrifugation, storage, and storage temperature, might have varied among the samples, affecting miRNA plasma levels (45, 46). However, confirmation of findings in both the study cohorts suggest a minimal impact of pre-analytical conditions in the results. Third, variations in the number of miRNAs identified in replicates of *in vitro* experiments may have led to the loss of some miRNAs. Fourth, the lack of tissue samples from organs with *Pf* sequestration restricted the histological confirmation of identified miRNAs, and the presence of co-infections other than blood culture positive bacteraemia cannot be neglected in the studied plasma samples. Finally, the association of each miRNA with specific symptoms that are part of the SM case definition may need further validation using a larger sample size, considering that our numbers were relatively small for individual SM criteria. In addition, future studies using the machine-learning approaches would allow the identification of a combination of miRNAs that may detect SM pathologies.

In conclusion, the profiling of miRNAs in media from HBE cells after incubation with a cytoadherent *Pf* strain and in plasmas from Mozambican children with different clinical presentations allowed the identification of promising miRNA candidates for characterizing severe malaria, specifically hsa-miR-4497. This study opens the ground for future analyses to understand the value of these miRNAs as a prognostic biomarker and for disentangling the aetiology of severe malaria.

## Data Availability

The datasets analysed in this study are available from the corresponding author on request.

## Acknowledgements

We are grateful to the children who participated in the study; the staff of the Manhiça District Hospital; the clinical officers, field supervisors and data managers; G. Cabrera, L. Mussacate, N. Ernesto José and A. Nhabomba for their contribution to the collection of parasites; L. Puyol for her laboratory management, as well as everyone who supported this study directly or indirectly. We also thank Ruhi Sikka, Varun Sharma, Rebecca Smith-Aguasca, Malia Skjefte, and Catriona Patterson for their useful comments on this manuscript.

^±^HG moved to the department of Infection Biology, London School of Hygiene and Tropical Medicine, London, UK.

This work was supported by the Instituto de Salud Carlos III (PI13/01478 cofunded by the Fondo Europeo de Desarrollo Regional [FEDER], CES10/021-I3SNS to AM and CP11/00269 from the Miguel Servet program to QB). HG was supported (Jan/2017 – Jan/2019) by the Science and Engineering Research Board (SERB), Department of Science & Technology, Government of India (Overseas Postdoctoral Fellowship, SB/OS/PDF-043/2015-16). ISGlobal is a member of the CERCA Programme, Generalitat de Catalunya (http://cerca.cat/en/suma/). CISM is supported by the Government of Mozambique and the Spanish Agency for International Development (AECID). This research is part of ISGlobal’s Program on the Molecular Mechanisms of Malaria, which is partially supported by the Fundación Ramón Areces.

## Biographical Sketch

Dr. Gupta is a molecular biologist and an early career malaria disease researcher. His research focuses on host and parasite factors associated with severe malaria, and on the use of molecular tools for the active surveillance of emerging drug resistance, gene deletions, and afebrile malaria in malaria-endemic regions.

## Availability of data and materials

The datasets analysed in this study are available from the corresponding author on request.

## Authors’ contributions

HG, MR carried out the molecular analysis, results interpretation and wrote the first draft of this manuscript. PC also carried out molecular analysis and conducted cytoadhesion assays. AS, RV, LM and IC participated in fieldwork, collected clinical, epidemiological data, plasma samples, dried blood drop filter papers and performed microscopy. AJ, XMV and DB participated in HRP2 analyse. MR, PC, HG, LP, AB and MB participated in bioinformatics and statistical analyses. QB and AM participated in the study design, supervision, funding acquisition, project administration and coordinated all the stages of the project. All authors reviewed and approved the final manuscript.

## Technical Appendix

### Material and Methods

#### Study population

Clinical malaria was defined as the presence of fever (axillary temperature≥37.5°) with an asexual parasitemia of *Pf* ≥500/µL. Children with severe malaria (SM) were those with at least one of the following symptoms: cerebral malaria (Blantyre Coma Score ≤2), severe anaemia (SA, packed cell volume <15% or haemoglobin <5g/dL), acidosis or acute respiratory distress (ARD, lactate >5mM and/or chest in-drawing or deep breathing), prostration (inability to sit or breastfeed in children old enough to do so based on their age), hypoglycaemia (blood glucose <2.2mM) and multiple seizures (≥2 convulsions in the preceding 24h) following the modified WHO criteria (1). Children with uncomplicated malaria (UM) were those with clinical malaria but not presenting any signs/symptoms of severity mentioned above (2). The presence of concomitant bacteraemia was tested in all SM cases using blood cultures and children with positive bacteraemia were excluded. Children with SM were treated according to Mozambican national guidelines with parenteral quinine in 2006 or parenteral artesunate (complemented with an oral artemisinin-based combination therapy) in 2014, and those with UM were treated with a combination of oral amodiaquine and sulfadoxine-pyrimethamine (Fansidar®) in 2006 or with artemether-lumefantrine (Coartem®) in 2014. Ten ml of heparinized blood was collected from study participants and processed within 2 hours after collection. Filter paper dried blood spots of 60µL blood were prepared from the vacutainer blood. After centrifugation at 1000rpms for 10minutes at 4°C, plasma was stored at - 20°C. The 2014 study was conducted as a quasi-exact repetition of the 2006 study, the only difference being that cases and controls were matched by parasitaemia level. Biochemistry parameters (glucose and lactate) and a full blood count were performed for each patient using Vitros DT60 and Sysmex Kx21 analyzers, respectively.

#### Parasitological determinations

Histidine-rich protein 2 (HRP2) levels were quantified using commercially available enzyme-linked immunosorbent assay kits (Malaria Ag CELISA; Cellabs Pty. Ltd., Brookvale, New South Wales, Australia) and an in-house highly sensitive quantitative bead suspension array based on Luminex technology. In brief, plasma samples were incubated overnight at 4°C with 2000 magnetic beads to a final dilution of 1:10. After washing, beads were sequentially incubated with 100 µL of in-house biotinylated antibody α-HRP2 (MBS832975, MyBiSource, San Diego, CL) at 1 µg/ml and with streptavidin-PE (42250-1ML, Sigma Aldrich, St. Louis, MO) at 1:1000 dilution. Finally, beads were washed and re-suspended in assay buffer, and the plate was read using the Luminex xMAP® 100/200 analyser (Luminex Corp., Austin, TX). A minimum of 50 microspheres per analyte were acquired and results were exported as crude median fluorescent intensity (MFI). Background (blank) MFIs were subtracted and normalized to account for plate to plate variation. Quantification was performed against a 5-parameter logistic regression curve fitted from a calibration curve consisting of recombinant protein HRP2 type A (890015, Microcoat GmbH, Germany).

#### *Pf* cytoadhesion assays

Human brain microvascular endothelial cells (Innoprot, Reference P10361) were cultured in 12 well-plates following the supplier’s recommendations and were left until 40% confluency was achieved. HBE cells were incubated in triplicate with *Pf*-iEs at trophozoite stage of the ePCR- binding FCR3 strain (ePCR-iE; which expresses the PfEPM1 protein that binds to ePCR receptor) and 3D7 strain (3D7-iE; a strain without the protein that binds to ePCR receptor) with 5% of both parasitemia and haematocrit. Non-infected erythrocytes were used as negative control. The cell- conditioned media of each group were collected after 1hr (t1) and 24hrs of stimulation (t24). Next day, HBE cells were stimulated as described above, after 1-hour incubation in agitation, cells were washed at least 10 times with binding media and fixed with 2% glutaraldehyde (SIGMA) in PBS (Gibco) overnight to assess adhesion by light microscopy. After washing with water, cells were stained with 10% Giemsa. iE and niE bound were counted in six different wells per assay in at least 500 nuclei cell per well. Results were presented as the number of adhered iE per 500 nuclei of cells. Estimation of *Pf* adhesion to purified receptors (CD36, CD54 and g1CqR) as well as platelet- mediated (PM)-agglutination and rosetting was performed as described elsewhere (2, 3).

Cytoadherence was defined as positive only if the number of iEs bound per mm^2^ was greater than the mean binding plus 2 standard deviations to Duffy-Fc coated petri dishes. *Pf* isolates were considered positive for PM clumping if the frequency of clumps was higher in the presence of platelets than in buffer-control and for rosetting if the frequency of rosettes was higher than 2% (2, 4).

#### Small RNA sequencing

Before RNA extraction, the level of haemolysis in plasma samples was assessed by spectrophotometry (EPOCH, BioTek) at a wavelength of 414nm (absorbance peak of free haemoglobin). Samples were classified as non-haemolysed if the optical density at 414nm was less than 0.2 (5). RNA was extracted from cell-conditioned media (3ml) and plasma samples (1ml) using the miRNeasy tissues/cells kit and miRNeasy Plasma/Serum kit (Qiagen), respectively, with the use of 5µg UltraPure™ glycogen/sample (Invitrogen). Given that the plasma samples were conserved in heparin, RNA was precipitated with LiCl as described elsewhere (6). Purified RNA quality and quantity were determined using the Bioanalyzer (Agilent Technologies) followed by preparation of libraries using NEBNext® Small RNA Library Prep Set for Illumina® (New England Biolabs), then separation of libraries in polyacrylamide gels (Novex, Invitrogen). The Bioanalyzer was again used to quantify and assess the size of the libraries. Further, libraries were pooled at the same equimolar concentrations and no more than 18 libraries were sequenced in the same lane using a HiSeq 2000 (Illumina) platform following the protocol for small RNAs (7).

A previously published pipeline was used to assess the sequencing quality, identification and quantification of small RNAs and normalization (7). First, a quality control (QC) was conducted using FASTX-Toolkit and FastQ Screen. After adaptor removing, reads with the following features were removed: 1) Reads <18nt, 2) Mean PHRED scores <30 and 3) Low complexity reads based on the mean score of the read. Good quality reads were then annotated to main RNA categories (tRNA, rRNA and miRNAs), and miRNA complexity was estimated as the number of distinct miRNAs that were observed in each sample. Finally, contamination with RNA from other species was evaluated by mapping reads to clade-specific mature miRNA sequences extracted from miRBase v21 (8). The tested species categories include animal sponges, nematodes, insects, lophotrochozoan, echinoderms, fish, birds, reptiles, rodents and primates.

Sequences that passed the QC were subjected to the seqBuster/seqCluster tool that retrieves miRNA and isomiRs counts (9, 10). To detect miRNAs and isomiRs, reads were mapped to the precursors and annotated to miRNAs or isomiRs using miRBase version 21 with the miraligner (9). DESeq2 R package v.1.10.1 (R version 3.3.2) (11) was used to perform an internal normalization where the counts for a miRNA in each sample were divided by the median of the ratios of observed counts to the geometric mean of each corresponding miRNAs over all samples.

#### Reverse transcriptase quantitative PCR

Fifty µl of plasma from the Mozambican children recruited in 2014 with no haemolysis were used for RNA extraction as described above. A synthetic RNA mimicking ath-miR-159a (*Arabidopsis thaliana*; Metabion) was added after lysis reaction at a final concentration of 1.5pM. cDNA synthesis and RT-qPCR [ABI PRISM 7500 HT Real-Time System (Applied Biosystems, Foster City, USA)] were performed using the TaqMan® Advanced miRNA assays. A standard curve of five serially diluted points was prepared with cDNA of six randomly selected samples and was run in triplicate for each miRNA. Results were normalized using a combination of endogenous controls (ECs). The selection of ECs was based on the following criteria: a) reported in scientific literature as previously used as ECs (12, 13), b) coefficient of variance (CV) of normalized counts across all samples ≤5%, c) basemean ≥3000, d) standard deviation ≤1 and e) log_2_fold change between SM and UM patients ≤1. Finally, the best two ECs tested as housekeepings using the NormFinder (14) were used for normalization of RT-qPCR data. miRNA relative expression levels (RELs) were calculated with the 2^−ΔCt^ method, where ΔCt = [Ct (miRNA) – Mean Ct (ECs)], considering efficiencies of 100% for all the miRNAs and ECs (12).

#### *In silico* analysis

The selected miRNAs were screened through four different gene target prediction programs: DIANA-microT-CDS (15), MiRDIP (16), MirGate (17), and TargetScan (http://www.targetscan.org/vert_71/). Identified gene targets of each program were compared using an online tool Venny2.1.0 (http://bioinfogp.cnb.csic.es/tools/venny/). The gene targets that occurred in more than one database were selected and screened through the miRTarBase (18) online program to check if these genes have been experimentally validated previously. These gene targets were anticipated to be true positive targets present at detectable levels in field samples. The identified gene targets were further analysed by DAVID 6.8 using *Homo sapiens* as the reference species. Genes were clustered to Gene Ontology terms and KEGG pathways (fold enrichment >1.5 and p <0.05).

### Statistical analysis

Differential expression of miRNAs and isomiRs was assessed using DESEq2 and IsomiRs packages in R (9, 10), which use negative binomial generalized linear models adjusted for multiple testing with the false discovery rate (FDR) by the Benjamini-Hochberg method (19). Those with an FDR of 5% or below were selected for posterior analysis. Analysis of the modification in the bases of the seed region was carried using isomiR package to determine a possible change in the target messenger RNAs. Mann-Whitney U and χ^2^ (Chi-square) tests were performed to compare continuous data and categorical data, respectively. Spearman correlation analysis was performed to assess the correlation of miRNA RELs (log transformed) with log transformed HRP2 levels. A two-sided p<0.05 was considered statistically significant. All statistical analyses were performed using R 3.3.2 in Linux-based system and graphs were prepared with GraphPad.

**Figure 1:**
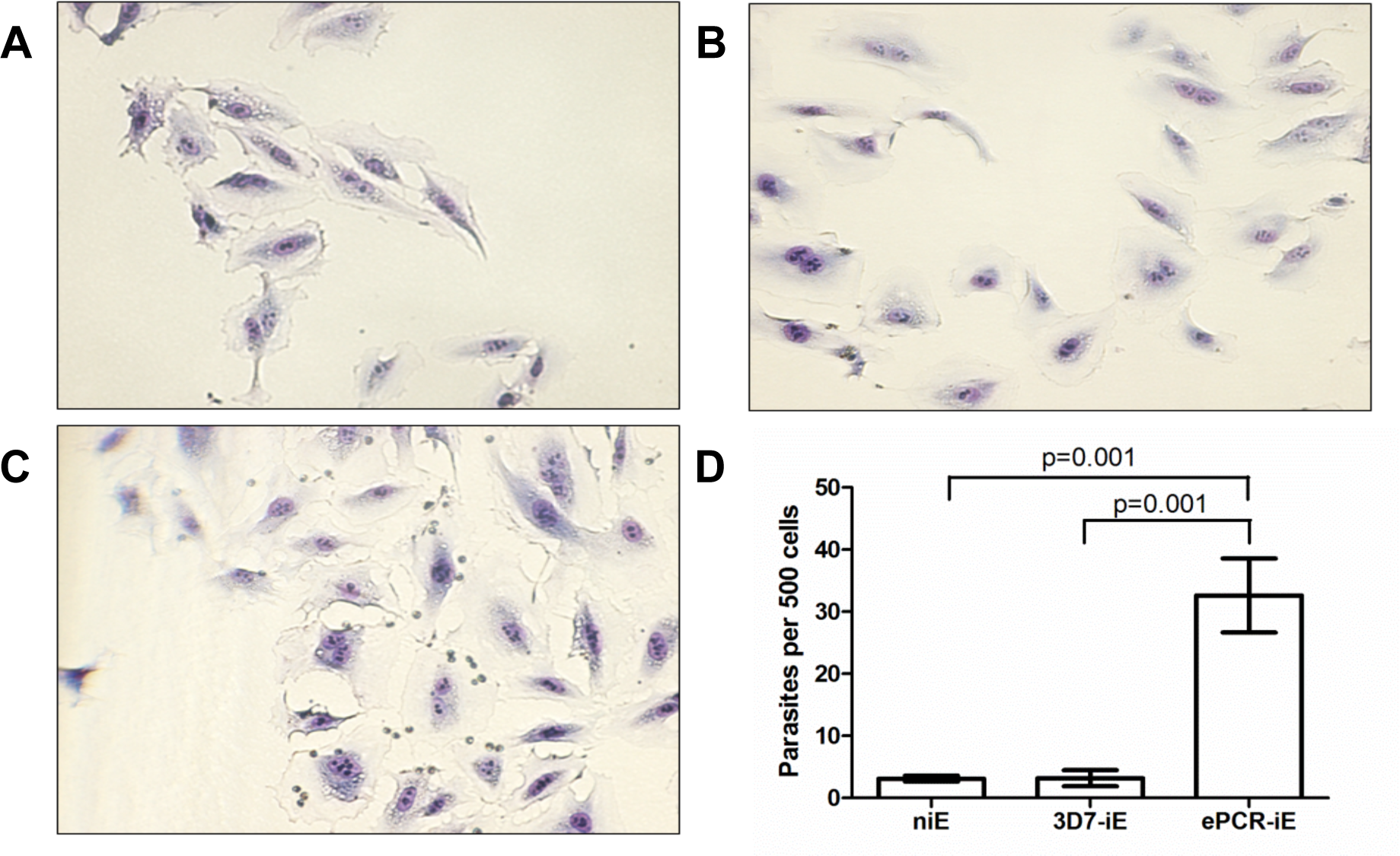
Cytoadhesion assay: Infected erythrocytes (iEs) with human brain endothelial (HBE) cells stained with Giemsa and visualised at 200X. A) non-infected erythrocytes (niE), B) non- adherent 3D7-infected erythrocytes (3D7-iE), C) adherent FCR3 expression endothelial receptor of protein C-infected erythrocytes (ePCR-iE) and D) three group’s comparison for infected erythrocytes adhered to HBE cells. Bars represent the mean and T line the standard deviation. p values were calculated using an unpaired t-test.

**Figure 2:**
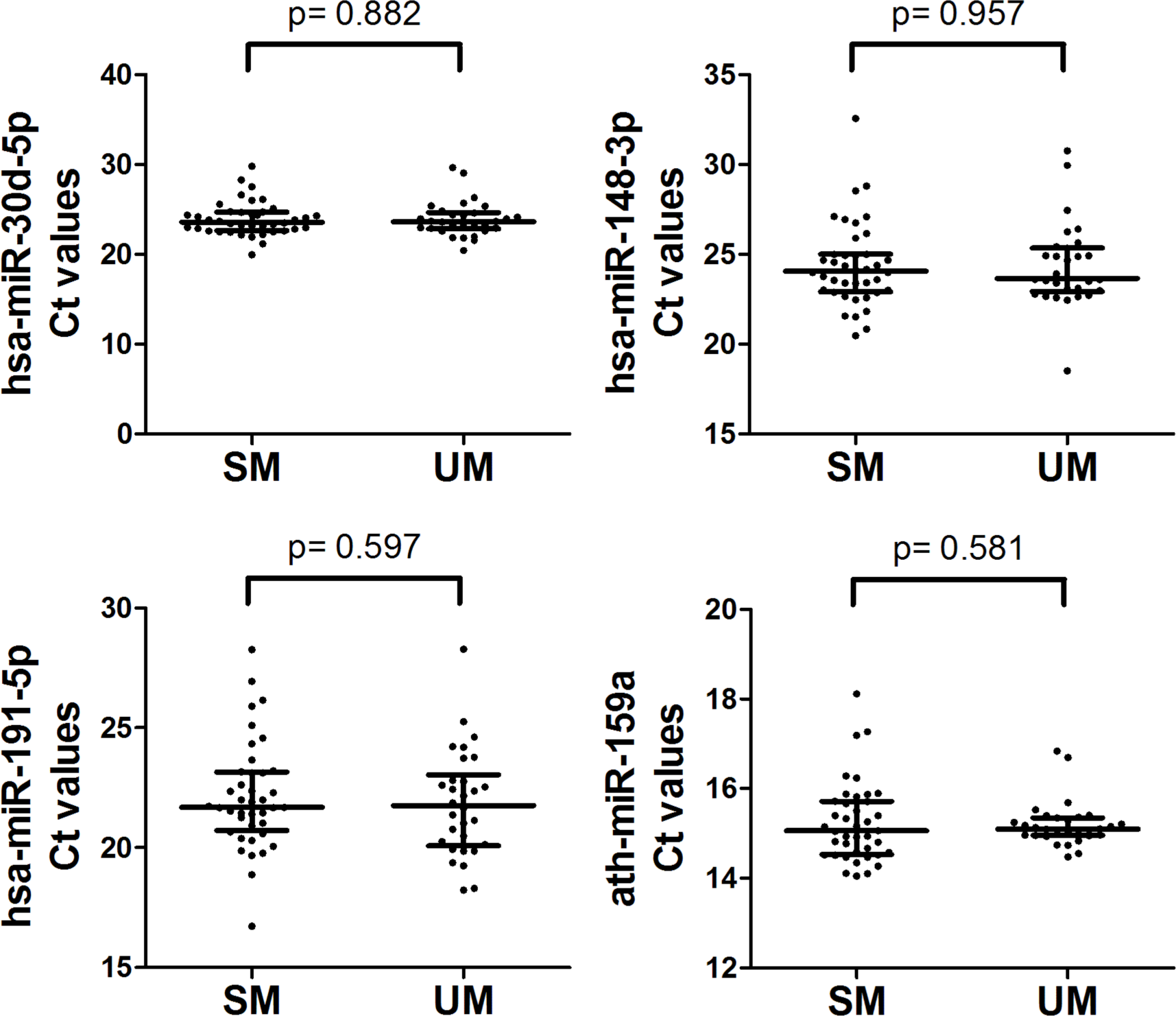
**RT-qPCR Ct values of exogenous (ath-miR-159a) and endogenous (hsa-miR-191-5p, hsa-miR-30d-5p and hsa-miR-148a-3p) controls in severe malaria (SM) and uncomplicated malaria (UM) groups.** Distributions were compared using Mann-Whitney U test. T bars represent median and Interquartile Ranges (IQR).

**Table 1:**
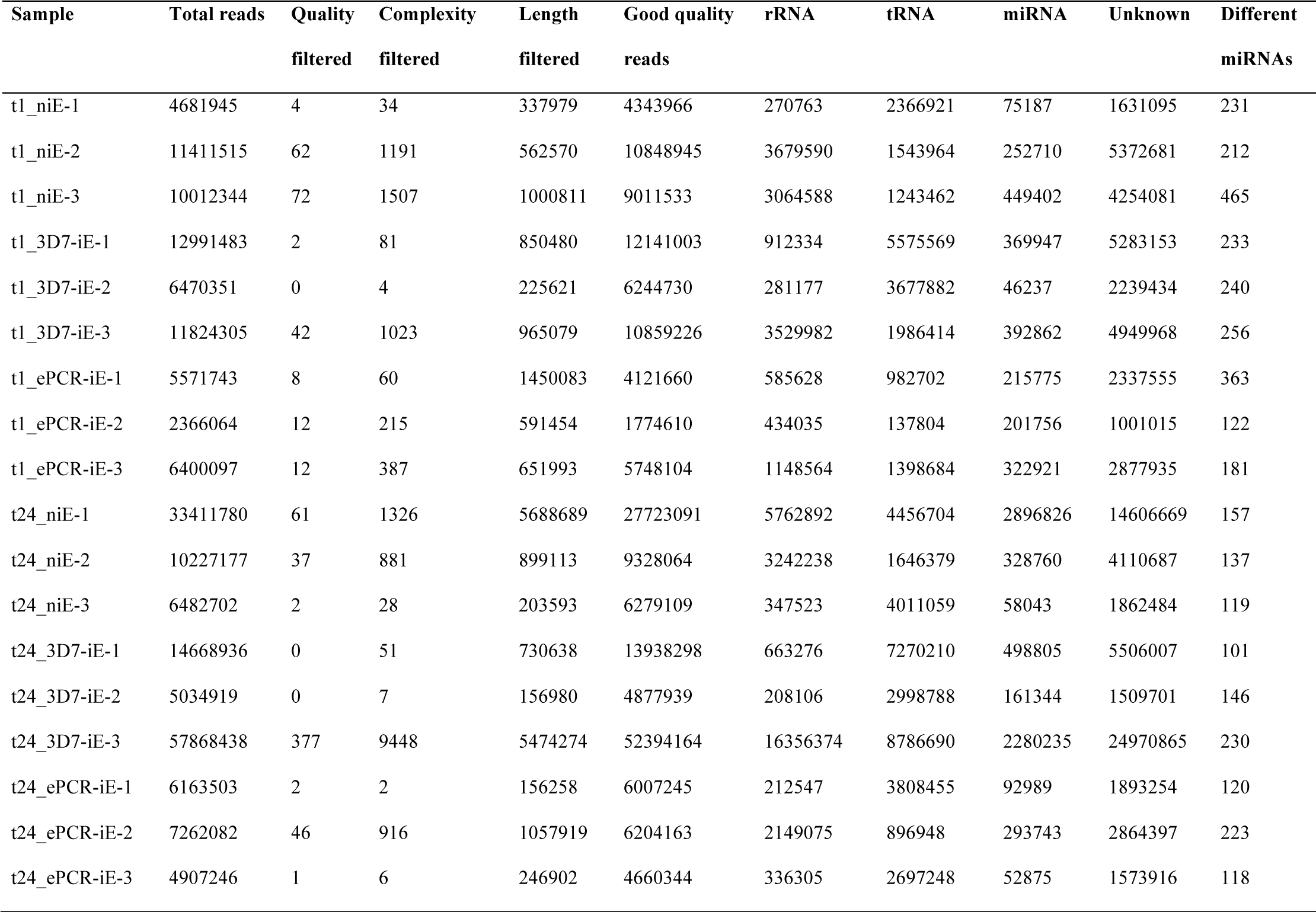
Number of reads, quality control and number of different miRNAs detected in cell-conditioned media of human brain endothelial cells exposed to cytoadherent (ePCR-iE), non-cytoadherent (3D7-iE) *P. falciparum* infected and non-infected erythrocytes (niE).

| Sample | Total reads | Quality<br>filtered | Complexity<br>filtered | Length<br>filtered | Good quality<br>reads | rRNA | tRNA | miRNA | Unknown | Different<br>miRNAs |
| --- | --- | --- | --- | --- | --- | --- | --- | --- | --- | --- |
| t1_niE-1 | 4681945 | 4 | 34 | 337979 | 4343966 | 270763 | 2366921 | 75187 | 1631095 | 231 |
| t1_niE-2 | 11411515 | 62 | 1191 | 562570 | 10848945 | 3679590 | 1543964 | 252710 | 5372681 | 212 |
| t1_niE-3 | 10012344 | 72 | 1507 | 1000811 | 9011533 | 3064588 | 1243462 | 449402 | 4254081 | 465 |
| t1_3D7-iE-1 | 12991483 | 2 | 81 | 850480 | 12141003 | 912334 | 5575569 | 369947 | 5283153 | 233 |
| t1_3D7-iE-2 | 6470351 | 0 | 4 | 225621 | 6244730 | 281177 | 3677882 | 46237 | 2239434 | 240 |
| t1_3D7-iE-3 | 11824305 | 42 | 1023 | 965079 | 10859226 | 3529982 | 1986414 | 392862 | 4949968 | 256 |
| t1_ePCR-iE-1 | 5571743 | 8 | 60 | 1450083 | 4121660 | 585628 | 982702 | 215775 | 2337555 | 363 |
| t1_ePCR-iE-2 | 2366064 | 12 | 215 | 591454 | 1774610 | 434035 | 137804 | 201756 | 1001015 | 122 |
| t1_ePCR-iE-3 | 6400097 | 12 | 387 | 651993 | 5748104 | 1148564 | 1398684 | 322921 | 2877935 | 181 |
| t24_niE-1 | 33411780 | 61 | 1326 | 5688689 | 27723091 | 5762892 | 4456704 | 2896826 | 14606669 | 157 |
| t24_niE-2 | 10227177 | 37 | 881 | 899113 | 9328064 | 3242238 | 1646379 | 328760 | 4110687 | 137 |
| t24_niE-3 | 6482702 | 2 | 28 | 203593 | 6279109 | 347523 | 4011059 | 58043 | 1862484 | 119 |
| t24_3D7-iE-1 | 14668936 | 0 | 51 | 730638 | 13938298 | 663276 | 7270210 | 498805 | 5506007 | 101 |
| t24_3D7-iE-2 | 5034919 | 0 | 7 | 156980 | 4877939 | 208106 | 2998788 | 161344 | 1509701 | 146 |
| t24_3D7-iE-3 | 57868438 | 377 | 9448 | 5474274 | 52394164 | 16356374 | 8786690 | 2280235 | 24970865 | 230 |
| t24_ePCR-iE-1 | 6163503 | 2 | 2 | 156258 | 6007245 | 212547 | 3808455 | 92989 | 1893254 | 120 |
| t24_ePCR-iE-2 | 7262082 | 46 | 916 | 1057919 | 6204163 | 2149075 | 896948 | 293743 | 2864397 | 223 |
| t24_ePCR-iE-3 | 4907246 | 1 | 6 | 246902 | 4660344 | 336305 | 2697248 | 52875 | 1573916 | 118 |

**Table 2:**
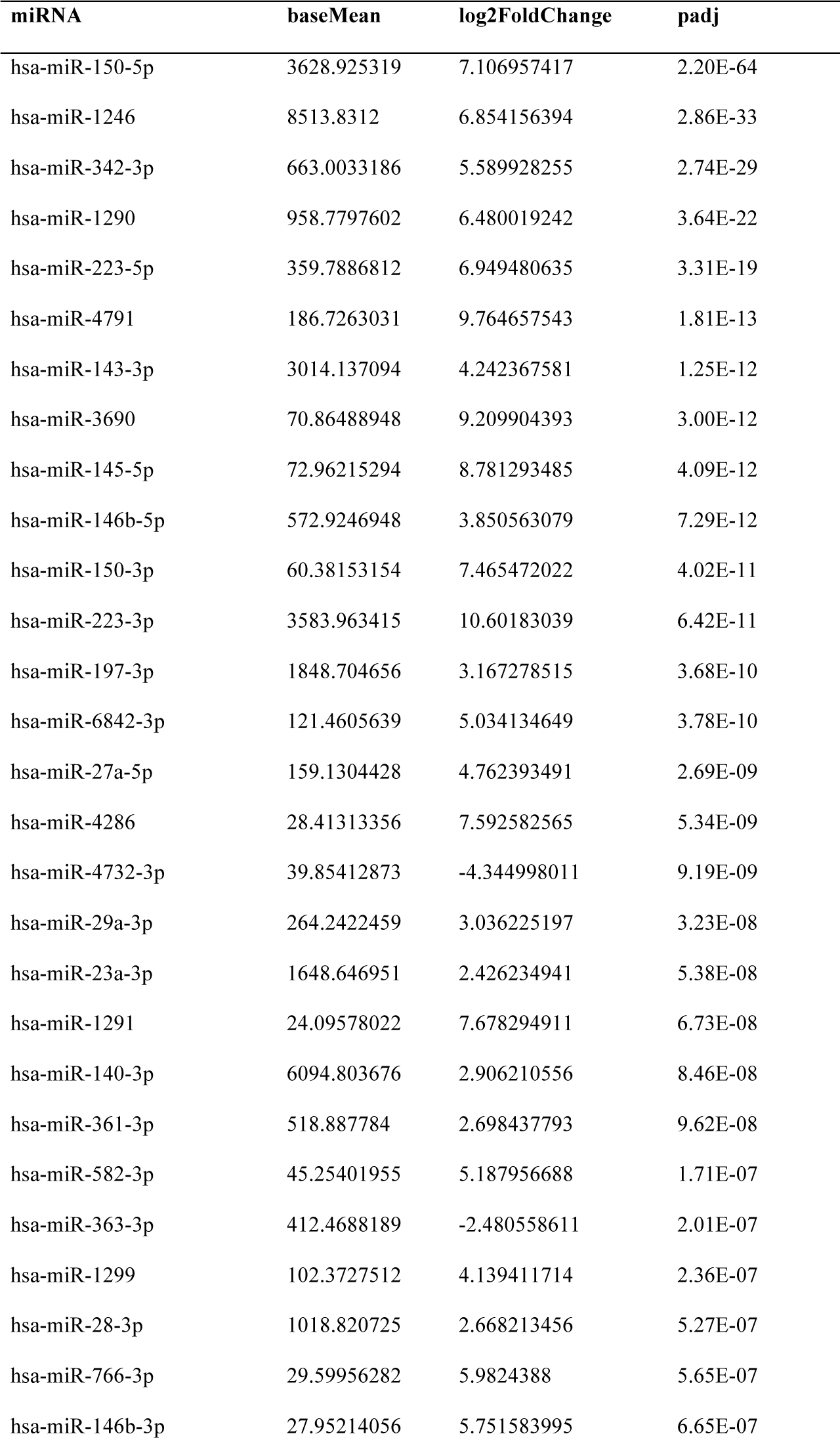
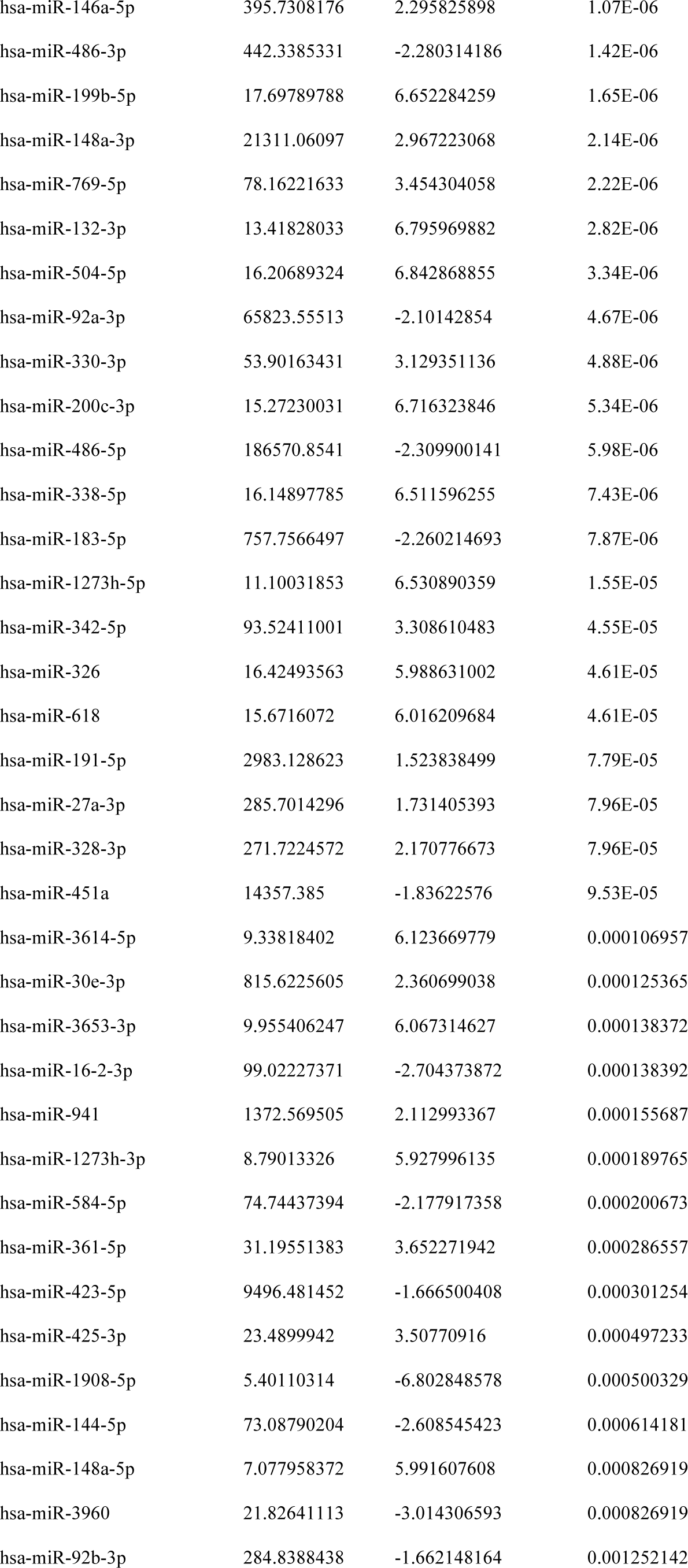
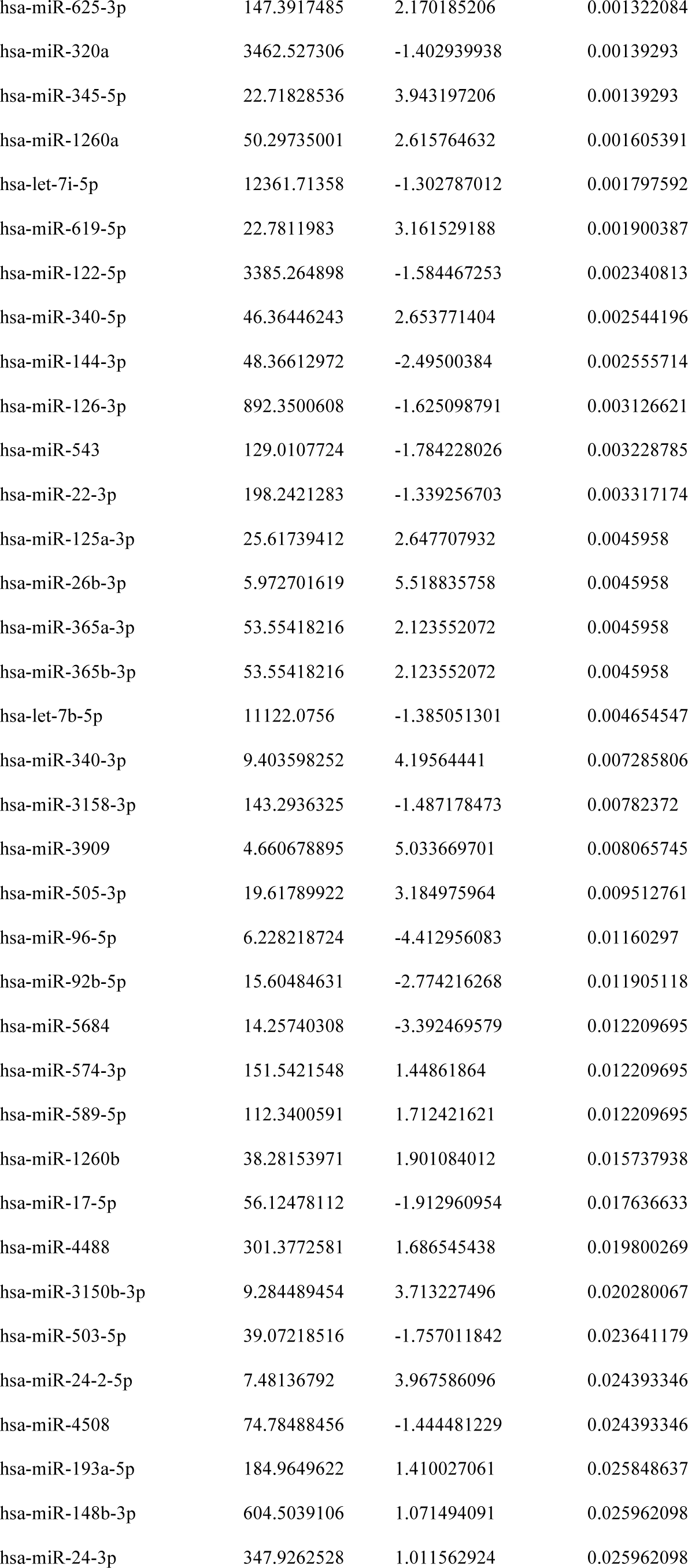
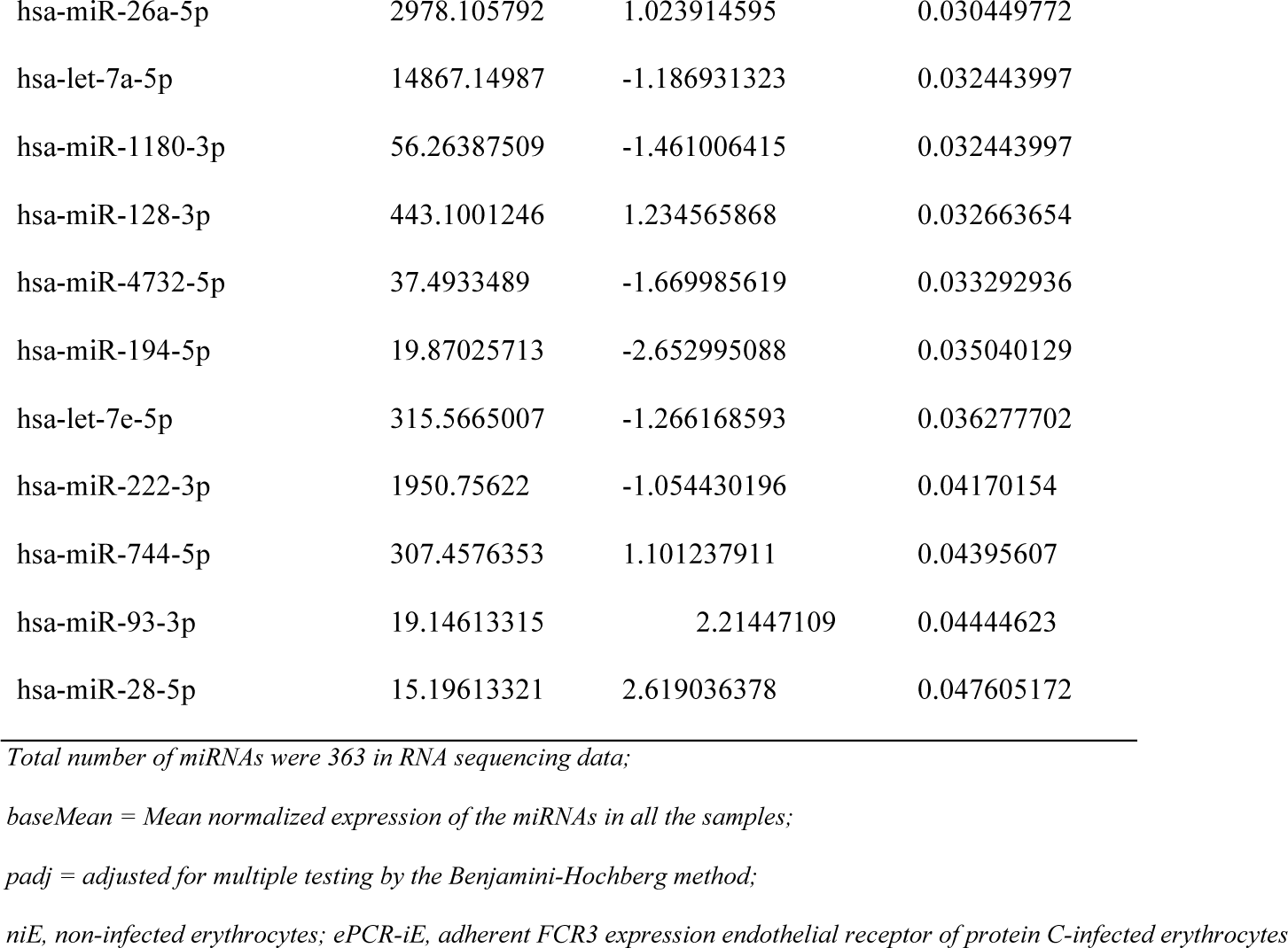
miRNAs differentially expressed in cell-conditioned media of human brain endothelial cells when exposed to niE and compared with ePCR-iE after one hour incubation. Positive FoldChange indicates overexpression in niE.

| miRNA | baseMean | log2FoldChange | padj |
| --- | --- | --- | --- |
| hsa-miR-150-5p | 3628.925319 | 7.106957417 | 2.20E-64 |
| hsa-miR-1246 | 8513.8312 | 6.854156394 | 2.86E-33 |
| hsa-miR-342-3p | 663.0033186 | 5.589928255 | 2.74E-29 |
| hsa-miR-1290 | 958.7797602 | 6.480019242 | 3.64E-22 |
| hsa-miR-223-5p | 359.7886812 | 6.949480635 | 3.31E-19 |
| hsa-miR-4791 | 186.7263031 | 9.764657543 | 1.81E-13 |
| hsa-miR-143-3p | 3014.137094 | 4.242367581 | 1.25E-12 |
| hsa-miR-3690 | 70.86488948 | 9.209904393 | 3.00E-12 |
| hsa-miR-145-5p | 72.96215294 | 8.781293485 | 4.09E-12 |
| hsa-miR-146b-5p | 572.9246948 | 3.850563079 | 7.29E-12 |
| hsa-miR-150-3p | 60.38153154 | 7.465472022 | 4.02E-11 |
| hsa-miR-223-3p | 3583.963415 | 10.60183039 | 6.42E-11 |
| hsa-miR-197-3p | 1848.704656 | 3.167278515 | 3.68E-10 |
| hsa-miR-6842-3p | 121.4605639 | 5.034134649 | 3.78E-10 |
| hsa-miR-27a-5p | 159.1304428 | 4.762393491 | 2.69E-09 |
| hsa-miR-4286 | 28.41313356 | 7.592582565 | 5.34E-09 |
| hsa-miR-4732-3p | 39.85412873 | -4.344998011 | 9.19E-09 |
| hsa-miR-29a-3p | 264.2422459 | 3.036225197 | 3.23E-08 |
| hsa-miR-23a-3p | 1648.646951 | 2.426234941 | 5.38E-08 |
| hsa-miR-1291 | 24.09578022 | 7.678294911 | 6.73E-08 |
| hsa-miR-140-3p | 6094.803676 | 2.906210556 | 8.46E-08 |
| hsa-miR-361-3p | 518.887784 | 2.698437793 | 9.62E-08 |
| hsa-miR-582-3p | 45.25401955 | 5.187956688 | 1.71E-07 |
| hsa-miR-363-3p | 412.4688189 | -2.480558611 | 2.01E-07 |
| hsa-miR-1299 | 102.3727512 | 4.139411714 | 2.36E-07 |
| hsa-miR-28-3p | 1018.820725 | 2.668213456 | 5.27E-07 |
| hsa-miR-766-3p | 29.59956282 | 5.9824388 | 5.65E-07 |
| hsa-miR-146b-3p | 27.95214056 | 5.751583995 | 6.65E-07 |
| hsa-miR-146a-5p | 395.7308176 | 2.295825898 | 1.07E-06 |
| hsa-miR-486-3p | 442.3385331 | -2.280314186 | 1.42E-06 |
| hsa-miR-199b-5p | 17.69789788 | 6.652284259 | 1.65E-06 |
| hsa-miR-148a-3p | 21311.06097 | 2.967223068 | 2.14E-06 |
| hsa-miR-769-5p | 78.16221633 | 3.454304058 | 2.22E-06 |
| hsa-miR-132-3p | 13.41828033 | 6.795969882 | 2.82E-06 |
| hsa-miR-504-5p | 16.20689324 | 6.842868855 | 3.34E-06 |
| hsa-miR-92a-3p | 65823.55513 | -2.10142854 | 4.67E-06 |
| hsa-miR-330-3p | 53.90163431 | 3.129351136 | 4.88E-06 |
| hsa-miR-200c-3p | 15.27230031 | 6.716323846 | 5.34E-06 |
| hsa-miR-486-5p | 186570.8541 | -2.309900141 | 5.98E-06 |
| hsa-miR-338-5p | 16.14897785 | 6.511596255 | 7.43E-06 |
| hsa-miR-183-5p | 757.7566497 | -2.260214693 | 7.87E-06 |
| hsa-miR-1273h-5p | 11.10031853 | 6.530890359 | 1.55E-05 |
| hsa-miR-342-5p | 93.52411001 | 3.308610483 | 4.55E-05 |
| hsa-miR-326 | 16.42493563 | 5.988631002 | 4.61E-05 |
| hsa-miR-618 | 15.6716072 | 6.016209684 | 4.61E-05 |
| hsa-miR-191-5p | 2983.128623 | 1.523838499 | 7.79E-05 |
| hsa-miR-27a-3p | 285.7014296 | 1.731405393 | 7.96E-05 |
| hsa-miR-328-3p | 271.7224572 | 2.170776673 | 7.96E-05 |
| hsa-miR-451a | 14357.385 | -1.83622576 | 9.53E-05 |
| hsa-miR-3614-5p | 9.33818402 | 6.123669779 | 0.000106957 |
| hsa-miR-30e-3p | 815.6225605 | 2.360699038 | 0.000125365 |
| hsa-miR-3653-3p | 9.955406247 | 6.067314627 | 0.000138372 |
| hsa-miR-16-2-3p | 99.02227371 | -2.704373872 | 0.000138392 |
| hsa-miR-941 | 1372.569505 | 2.112993367 | 0.000155687 |
| hsa-miR-1273h-3p | 8.79013326 | 5.927996135 | 0.000189765 |
| hsa-miR-584-5p | 74.74437394 | -2.177917358 | 0.000200673 |
| hsa-miR-361-5p | 31.19551383 | 3.652271942 | 0.000286557 |
| hsa-miR-423-5p | 9496.481452 | -1.666500408 | 0.000301254 |
| hsa-miR-425-3p | 23.4899942 | 3.50770916 | 0.000497233 |
| hsa-miR-1908-5p | 5.40110314 | -6.802848578 | 0.000500329 |
| hsa-miR-144-5p | 73.08790204 | -2.608545423 | 0.000614181 |
| hsa-miR-148a-5p | 7.077958372 | 5.991607608 | 0.000826919 |
| hsa-miR-3960 | 21.82641113 | -3.014306593 | 0.000826919 |
| hsa-miR-92b-3p | 284.8388438 | -1.662148164 | 0.001252142 |
| hsa-miR-625-3p | 147.3917485 | 2.170185206 | 0.001322084 |
| hsa-miR-320a | 3462.527306 | -1.402939938 | 0.00139293 |
| hsa-miR-345-5p | 22.71828536 | 3.943197206 | 0.00139293 |
| hsa-miR-1260a | 50.29735001 | 2.615764632 | 0.001605391 |
| hsa-let-7i-5p | 12361.71358 | -1.302787012 | 0.001797592 |
| hsa-miR-619-5p | 22.7811983 | 3.161529188 | 0.001900387 |
| hsa-miR-122-5p | 3385.264898 | -1.584467253 | 0.002340813 |
| hsa-miR-340-5p | 46.36446243 | 2.653771404 | 0.002544196 |
| hsa-miR-144-3p | 48.36612972 | -2.49500384 | 0.002555714 |
| hsa-miR-126-3p | 892.3500608 | -1.625098791 | 0.003126621 |
| hsa-miR-543 | 129.0107724 | -1.784228026 | 0.003228785 |
| hsa-miR-22-3p | 198.2421283 | -1.339256703 | 0.003317174 |
| hsa-miR-125a-3p | 25.61739412 | 2.647707932 | 0.0045958 |
| hsa-miR-26b-3p | 5.972701619 | 5.518835758 | 0.0045958 |
| hsa-miR-365a-3p | 53.55418216 | 2.123552072 | 0.0045958 |
| hsa-miR-365b-3p | 53.55418216 | 2.123552072 | 0.0045958 |
| hsa-let-7b-5p | 11122.0756 | -1.385051301 | 0.004654547 |
| hsa-miR-340-3p | 9.403598252 | 4.19564441 | 0.007285806 |
| hsa-miR-3158-3p | 143.2936325 | -1.487178473 | 0.00782372 |
| hsa-miR-3909 | 4.660678895 | 5.033669701 | 0.008065745 |
| hsa-miR-505-3p | 19.61789922 | 3.184975964 | 0.009512761 |
| hsa-miR-96-5p | 6.228218724 | -4.412956083 | 0.01160297 |
| hsa-miR-92b-5p | 15.60484631 | -2.774216268 | 0.011905118 |
| hsa-miR-5684 | 14.25740308 | -3.392469579 | 0.012209695 |
| hsa-miR-574-3p | 151.5421548 | 1.44861864 | 0.012209695 |
| hsa-miR-589-5p | 112.3400591 | 1.712421621 | 0.012209695 |
| hsa-miR-1260b | 38.28153971 | 1.901084012 | 0.015737938 |
| hsa-miR-17-5p | 56.12478112 | -1.912960954 | 0.017636633 |
| hsa-miR-4488 | 301.3772581 | 1.686545438 | 0.019800269 |
| hsa-miR-3150b-3p | 9.284489454 | 3.713227496 | 0.020280067 |
| hsa-miR-503-5p | 39.07218516 | -1.757011842 | 0.023641179 |
| hsa-miR-24-2-5p | 7.48136792 | 3.967586096 | 0.024393346 |
| hsa-miR-4508 | 74.78488456 | -1.444481229 | 0.024393346 |
| hsa-miR-193a-5p | 184.9649622 | 1.410027061 | 0.025848637 |
| hsa-miR-148b-3p | 604.5039106 | 1.071494091 | 0.025962098 |
| hsa-miR-24-3p | 347.9262528 | 1.011562924 | 0.025962098 |
| hsa-miR-26a-5p | 2978.105792 | 1.023914595 | 0.030449772 |
| hsa-let-7a-5p | 14867.14987 | -1.186931323 | 0.032443997 |
| hsa-miR-1180-3p | 56.26387509 | -1.461006415 | 0.032443997 |
| hsa-miR-128-3p | 443.1001246 | 1.234565868 | 0.032663654 |
| hsa-miR-4732-5p | 37.4933489 | -1.669985619 | 0.033292936 |
| hsa-miR-194-5p | 19.87025713 | -2.652995088 | 0.035040129 |
| hsa-let-7e-5p | 315.5665007 | -1.266168593 | 0.036277702 |
| hsa-miR-222-3p | 1950.75622 | -1.054430196 | 0.04170154 |
| hsa-miR-744-5p | 307.4576353 | 1.101237911 | 0.04395607 |
| hsa-miR-93-3p | 19.14613315 | 2.21447109 | 0.04444623 |
| hsa-miR-28-5p | 15.19613321 | 2.619036378 | 0.047605172 |

*Total number of miRNAs were 363 in RNA sequencing data;*
*baseMean = Mean normalized expression of the miRNAs in all the samples;*
*padj = adjusted for multiple testing by the Benjamini-Hochberg method;*
*niE, non-infected erythrocytes; ePCR-iE, adherent FCR3 expression endothelial receptor of protein C-infected erythrocytes*

**Table 3:** miRNAs differentially expressed in cell-conditioned media of human brain endothelial cells when exposed to 3D7-iE and compared with ePCR-iE after one hour incubation. Positive FoldChange indicates overexpression in 3D7-iE.

| miRNA | baseMean | log2FoldChange | padj |
| --- | --- | --- | --- |
| hsa-miR-150-5p | 3628.925319 | 6.447621237 | 8.41E-53 |
| hsa-miR-1246 | 8513.8312 | 6.448500505 | 2.44E-29 |
| hsa-miR-342-3p | 663.0033186 | 4.805719054 | 1.87E-21 |
| hsa-miR-1290 | 958.7797602 | 6.047501723 | 3.28E-19 |
| hsa-miR-143-3p | 3014.137094 | 4.973540967 | 3.06E-17 |
| hsa-miR-223-5p | 359.7886812 | 6.431781207 | 1.96E-16 |
| hsa-miR-4791 | 186.7263031 | 10.38932761 | 2.56E-15 |
| hsa-miR-145-5p | 72.96215294 | 8.773111499 | 5.33E-12 |
| hsa-miR-23a-3p | 1648.646951 | 2.919080408 | 3.83E-11 |
| hsa-miR-146b-5p | 572.9246948 | 3.720217317 | 5.04E-11 |
| hsa-miR-423-5p | 9496.481452 | -2.845800299 | 1.45E-10 |
| hsa-miR-223-3p | 3583.963415 | 10.40824443 | 1.77E-10 |
| hsa-miR-28-3p | 1018.820725 | 3.236927509 | 7.54E-10 |
| hsa-miR-197-3p | 1848.704656 | 3.045327492 | 2.21E-09 |
| hsa-miR-3690 | 70.86488948 | 7.939040485 | 3.04E-09 |
| hsa-miR-363-3p | 412.4688189 | -2.752949366 | 7.66E-09 |
| hsa-miR-451a | 14357.385 | -2.623190083 | 1.10E-08 |
| hsa-miR-4732-3p | 39.85412873 | -4.446862598 | 1.22E-08 |
| hsa-miR-4286 | 28.41313356 | 7.221117949 | 3.72E-08 |
| hsa-miR-1299 | 102.3727512 | 4.296659106 | 8.94E-08 |
| hsa-miR-146a-5p | 395.7308176 | 2.416585365 | 3.27E-07 |
| hsa-miR-320a | 3462.527306 | -2.147110555 | 4.06E-07 |
| hsa-miR-150-3p | 60.38153154 | 5.841471731 | 5.53E-07 |
| hsa-miR-3158-3p | 143.2936325 | -2.671376413 | 6.03E-07 |
| hsa-miR-199b-5p | 17.69789788 | 6.756414418 | 1.41E-06 |
| hsa-miR-326 | 16.42493563 | 7.000378659 | 1.46E-06 |
| hsa-miR-92a-3p | 65823.55513 | -2.206241834 | 1.80E-06 |
| hsa-miR-618 | 15.6716072 | 6.882512894 | 2.54E-06 |
| hsa-miR-27a-5p | 159.1304428 | 3.853017578 | 2.61E-06 |
| hsa-miR-361-5p | 31.19551383 | 4.557109213 | 3.99E-06 |
| hsa-miR-766-3p | 29.59956282 | 5.475435141 | 7.17E-06 |
| hsa-miR-338-5p | 16.14897785 | 6.585842515 | 7.56E-06 |
| hsa-miR-1180-3p | 56.26387509 | -2.865258808 | 1.15E-05 |
| hsa-miR-486-5p | 186570.8541 | -2.257048303 | 1.36E-05 |
| hsa-miR-486-3p | 442.3385331 | -2.077360932 | 1.53E-05 |
| hsa-miR-140-3p | 6094.803676 | 2.378049094 | 1.89E-05 |
| hsa-miR-1291 | 24.09578022 | 6.218174392 | 2.27E-05 |
| hsa-miR-29a-3p | 264.2422459 | 2.377846317 | 2.52E-05 |
| hsa-miR-4508 | 74.78488456 | -2.537316275 | 3.24E-05 |
| hsa-miR-6842-3p | 121.4605639 | 3.458457239 | 3.89E-05 |
| hsa-miR-200c-3p | 15.27230031 | 6.110652566 | 5.42E-05 |
| hsa-miR-504-5p | 16.20689324 | 6.10449083 | 5.42E-05 |
| hsa-miR-582-3p | 45.25401955 | 4.02067135 | 9.79E-05 |
| hsa-miR-769-5p | 78.16221633 | 2.900427661 | 0.000122239 |
| hsa-miR-27a-3p | 285.7014296 | 1.699710038 | 0.000132728 |
| hsa-miR-625-3p | 147.3917485 | 2.549490741 | 0.000132728 |
| hsa-miR-340-5p | 46.36446243 | 3.201883577 | 0.000214609 |
| hsa-miR-144-3p | 48.36612972 | -3.0660485 | 0.000220627 |
| hsa-miR-425-3p | 23.4899942 | 3.724522238 | 0.000223367 |
| hsa-miR-320b | 420.1406035 | -1.862313231 | 0.000276457 |
| hsa-miR-365a-3p | 53.55418216 | 2.655154806 | 0.000276457 |
| hsa-miR-365b-3p | 53.55418216 | 2.655154806 | 0.000276457 |
| hsa-miR-132-3p | 13.41828033 | 5.410556286 | 0.000380553 |
| hsa-let-7i-5p | 12361.71358 | -1.47141141 | 0.000382512 |
| hsa-miR-30e-3p | 815.6225605 | 2.226049992 | 0.000382512 |
| hsa-miR-92b-5p | 15.60484631 | -4.163857943 | 0.0003924 |
| hsa-miR-146b-3p | 27.95214056 | 4.242630881 | 0.000482005 |
| hsa-miR-92b-3p | 284.8388438 | -1.785328471 | 0.000505668 |
| hsa-miR-4732-5p | 37.4933489 | -2.594968757 | 0.000643162 |
| hsa-miR-122-5p | 3385.264898 | -1.753744088 | 0.000704175 |
| hsa-miR-144-5p | 73.08790204 | -2.558810315 | 0.00089817 |
| hsa-miR-3653-3p | 9.955406247 | 5.420823215 | 0.000963768 |
| hsa-miR-330-3p | 53.90163431 | 2.33736255 | 0.001216256 |
| hsa-miR-2115-3p | 6.041154984 | 5.834660721 | 0.001395787 |
| hsa-miR-183-5p | 757.7566497 | -1.689145385 | 0.001439809 |
| hsa-miR-1273h-3p | 8.79013326 | 5.232979185 | 0.001476418 |
| hsa-miR-1273h-5p | 11.10031853 | 5.004655278 | 0.001837808 |
| hsa-miR-345-5p | 22.71828536 | 3.888926893 | 0.001844401 |
| hsa-miR-361-3p | 518.887784 | 1.669833063 | 0.001878529 |
| hsa-miR-328-3p | 271.7224572 | 1.756856697 | 0.002066471 |
| hsa-miR-3614-5p | 9.33818402 | 5.084630328 | 0.002066471 |
| hsa-miR-1260a | 50.29735001 | 2.56883531 | 0.002146885 |
| hsa-miR-191-5p | 2983.128623 | 1.22004135 | 0.002216696 |
| hsa-miR-505-3p | 19.61789922 | 3.426913927 | 0.00528521 |
| hsa-miR-4685-3p | 26.37952361 | -2.212963535 | 0.006078573 |
| hsa-miR-148a-3p | 21311.06097 | 1.831991607 | 0.006860876 |
| hsa-miR-126-3p | 892.3500608 | -1.523444179 | 0.007103832 |
| hsa-let-7b-5p | 11122.0756 | -1.342668008 | 0.007601427 |
| hsa-miR-5189-5p | 4.690302347 | -6.195706422 | 0.008125759 |
| hsa-miR-4448 | 19.99074734 | 2.751420355 | 0.008596921 |
| hsa-miR-28-5p | 15.19613321 | 3.329553964 | 0.008697526 |
| hsa-miR-619-5p | 22.7811983 | 2.721712643 | 0.011008689 |
| hsa-miR-5684 | 14.25740308 | -3.504790338 | 0.011818106 |
| hsa-miR-221-5p | 30.33237403 | 2.235527273 | 0.014993794 |
| hsa-miR-24-3p | 347.9262528 | 1.101827408 | 0.015833224 |
| hsa-miR-193a-5p | 184.9649622 | 1.491052572 | 0.020130112 |
| hsa-miR-22-3p | 198.2421283 | -1.113442482 | 0.021327869 |
| hsa-miR-93-3p | 19.14613315 | 2.52716344 | 0.021327869 |
| hsa-miR-2110 | 117.7715931 | -1.410612056 | 0.02301949 |
| hsa-miR-342-5p | 93.52411001 | 2.045398548 | 0.02301949 |
| hsa-miR-543 | 129.0107724 | -1.437070341 | 0.02604373 |
| hsa-miR-155-5p | 135.1369051 | 1.308204109 | 0.02781228 |
| hsa-miR-152-3p | 52.85362465 | 1.703147513 | 0.027818637 |
| hsa-miR-584-5p | 74.74437394 | -1.39071939 | 0.029085671 |
| hsa-let-7e-5p | 315.5665007 | -1.333615616 | 0.030343738 |
| hsa-miR-1908-5p | 5.40110314 | -4.518938959 | 0.033717462 |
| hsa-miR-16-2-3p | 99.02227371 | -1.642902948 | 0.035497673 |
| hsa-miR-3150b-3p | 9.284489454 | 3.477104992 | 0.037288673 |
| hsa-miR-106a-5p | 5.273323169 | -4.723234986 | 0.046728731 |
| hsa-miR-15b-5p | 59.88239009 | 1.339085366 | 0.048947187 |

*Total number of miRNAs were 363 in RNA sequencing data;*
*baseMean = Mean normalized expression of the miRNAs in all the samples;*
*padj = adjusted for multiple testing by the Benjamini-Hochberg method;*
*3D7-iE, non-adherent 3D7-infected erythrocytes; ePCR-iE, adherent FCR3 expression endothelial receptor of protein C-infected erythrocytes*

**Table 4:**
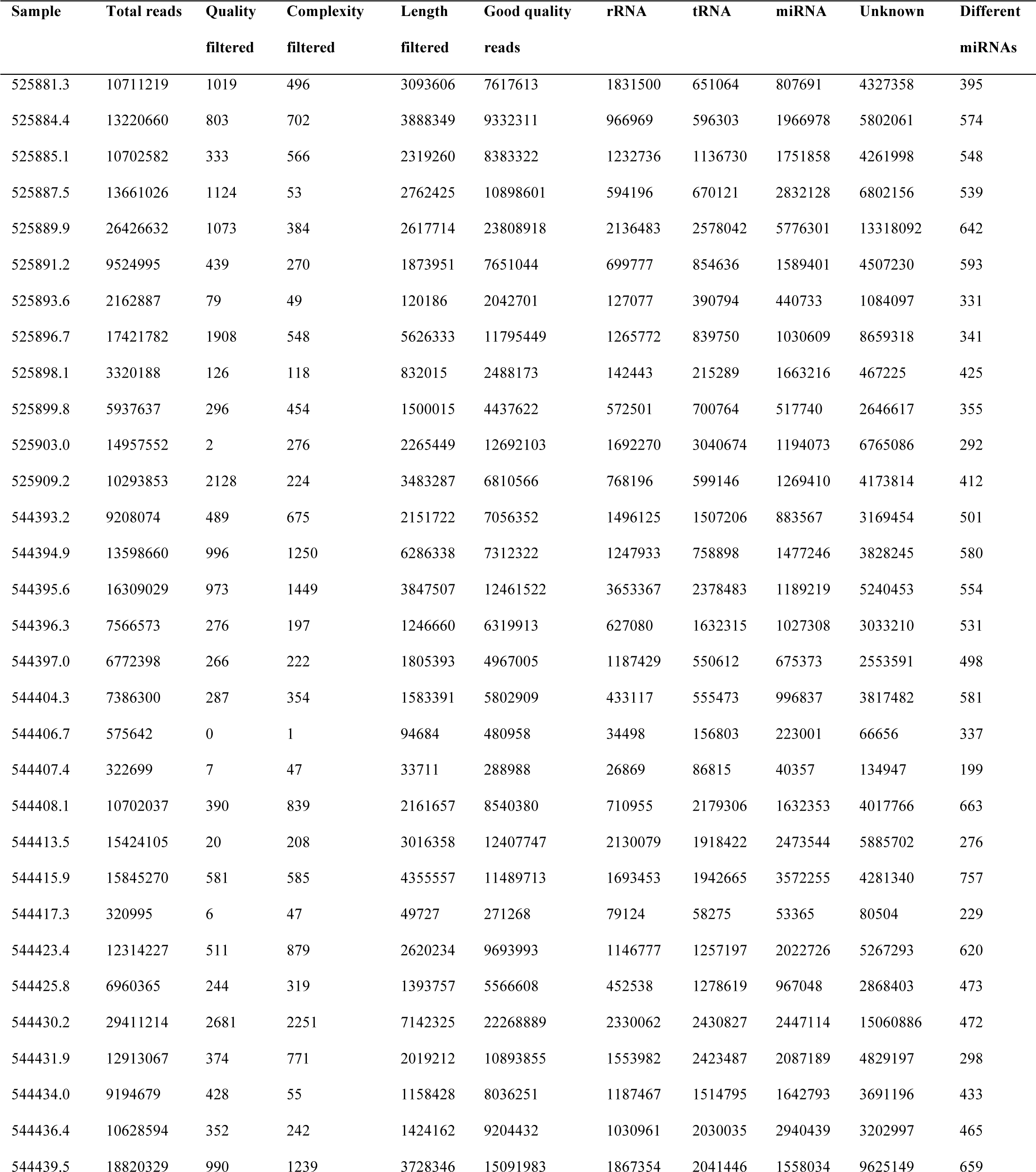
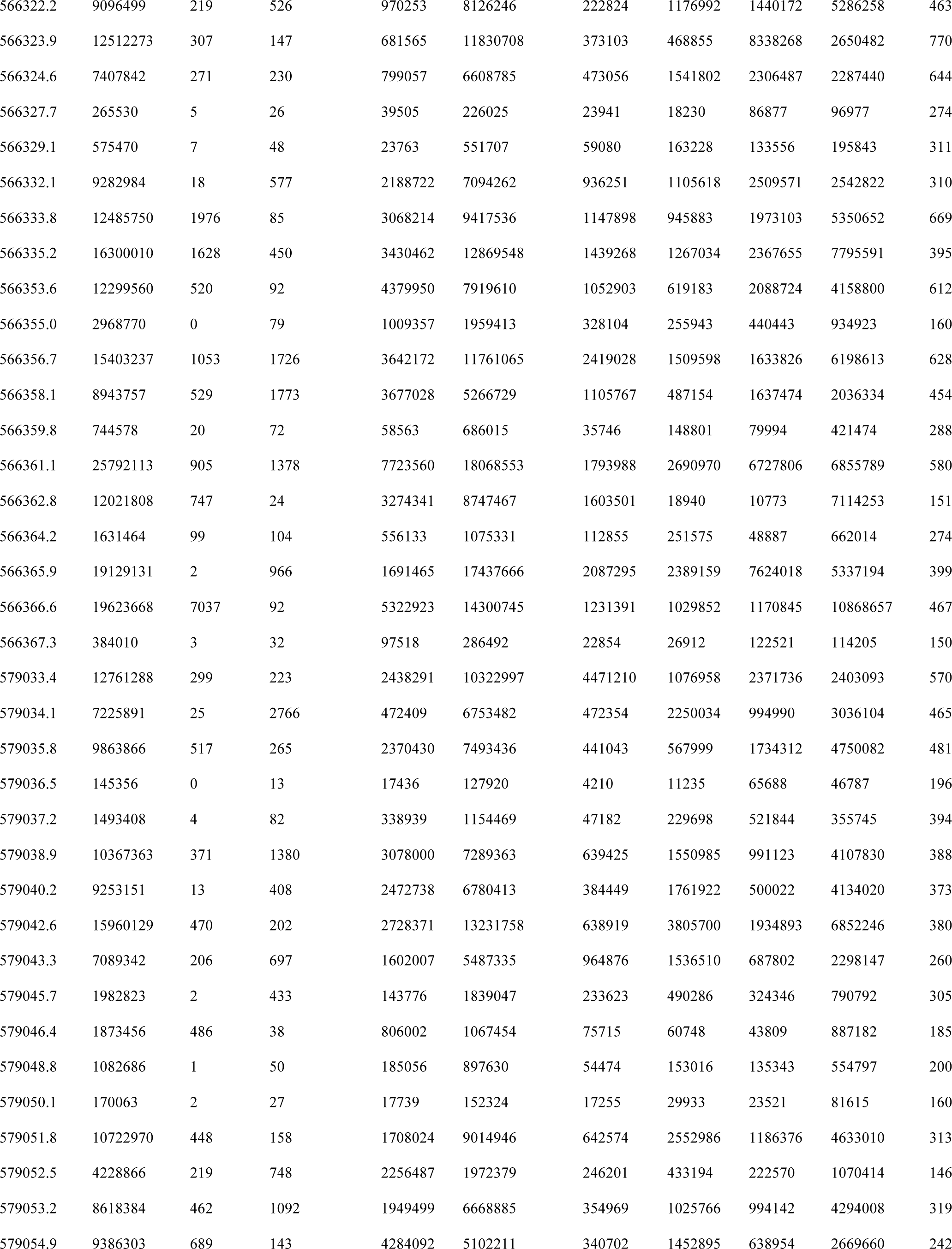
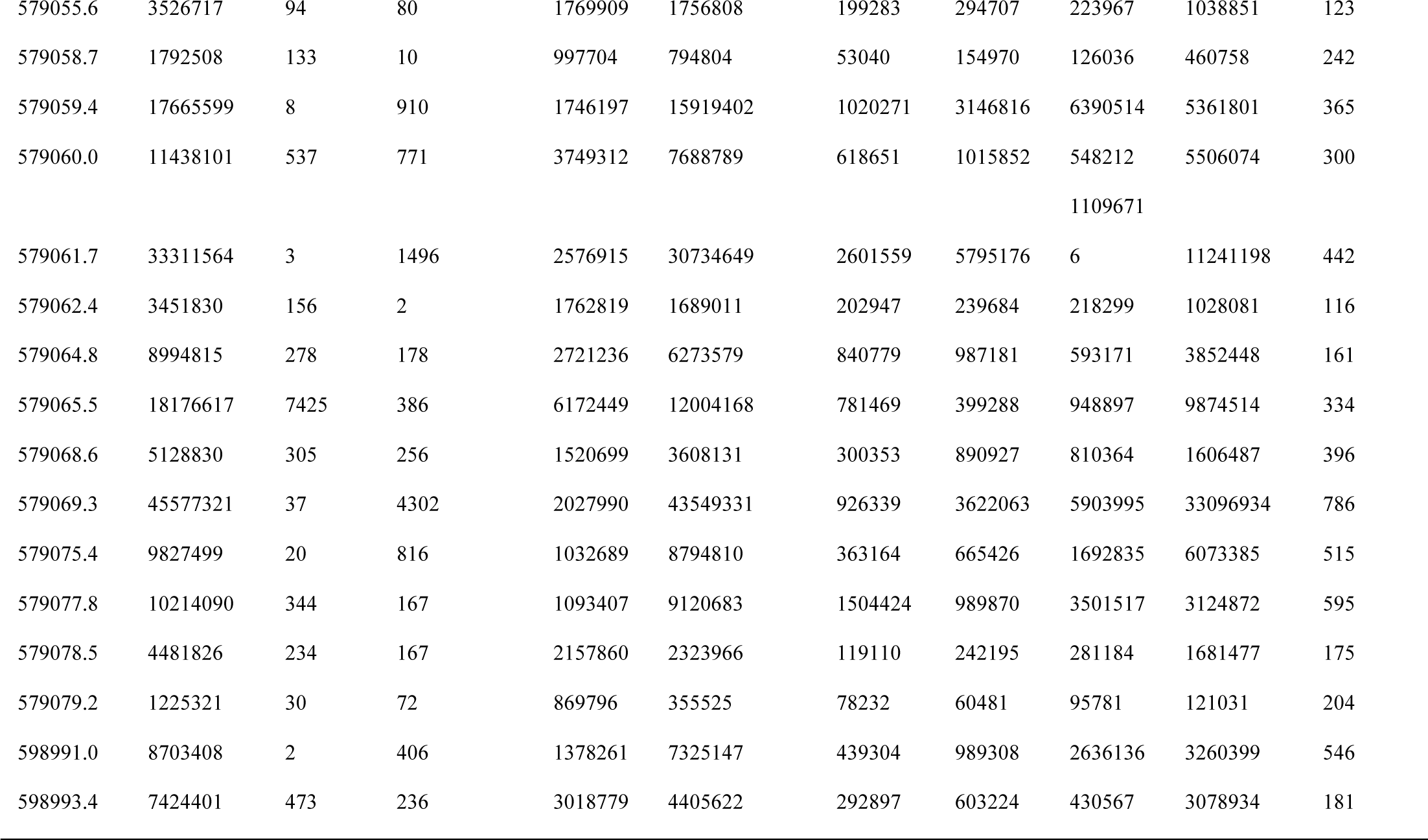
Number of reads, quality control and number of different miRNAs detected in plasma from Mozambican children recruited in 2006.

**Table 5:** Spearman correlations between ELISA based HRP2 levels and miRNA relative expression levels (RELs) in plasma samples from Mozambican children. HRP2 levels and miRNA RELs were log transformed.

| miRNA | rho | p value |
| --- | --- | --- |
| **Children recruited in 2006** |  |  |
| hsa-miR-10b-5p | 0.415 | 0.020 |
| hsa-miR-378a-3p | 0.422 | 0.020 |
| hsa-miR-4497 | 0.533 | <0.001 |
| **Children recruited in 2014** |  |  |
| hsa-miR-122-5p | -0.016 | 0.892 |
| hsa-miR-320a | 0.121 | 0.320 |
| hsa-miR-1246 | 0.066 | 0.588 |
| hsa-miR-1290 | 0.123 | 0.310 |
| hsa-miR-3158-3p | 0.511 | <0.001 |
| hsa-miR-4497 | 0.401 | <0.001 |

**Table 6:** PCR efficiencies of each miRNA used for RT-qPCR analysis.

| miRNA | PCR efficiencies |
| --- | --- |
| hsa-miR-122-5p | 99.6% |
| hsa-miR-320a | 95.3% |
| hsa-miR-1246 | 98.4% |
| hsa-miR-1290 | 103.0% |
| hsa-miR-3158-3p | 91.2% |
| hsa-miR-4497 | 96.9% |
| has-miR-191-5p | 93.1% |
| hsa-miR-30d-5p | 103.8% |
| hsa-miR-148a-3p | 100.1% |
| ath-miR-159a | 103.2% |

**Table 7:**
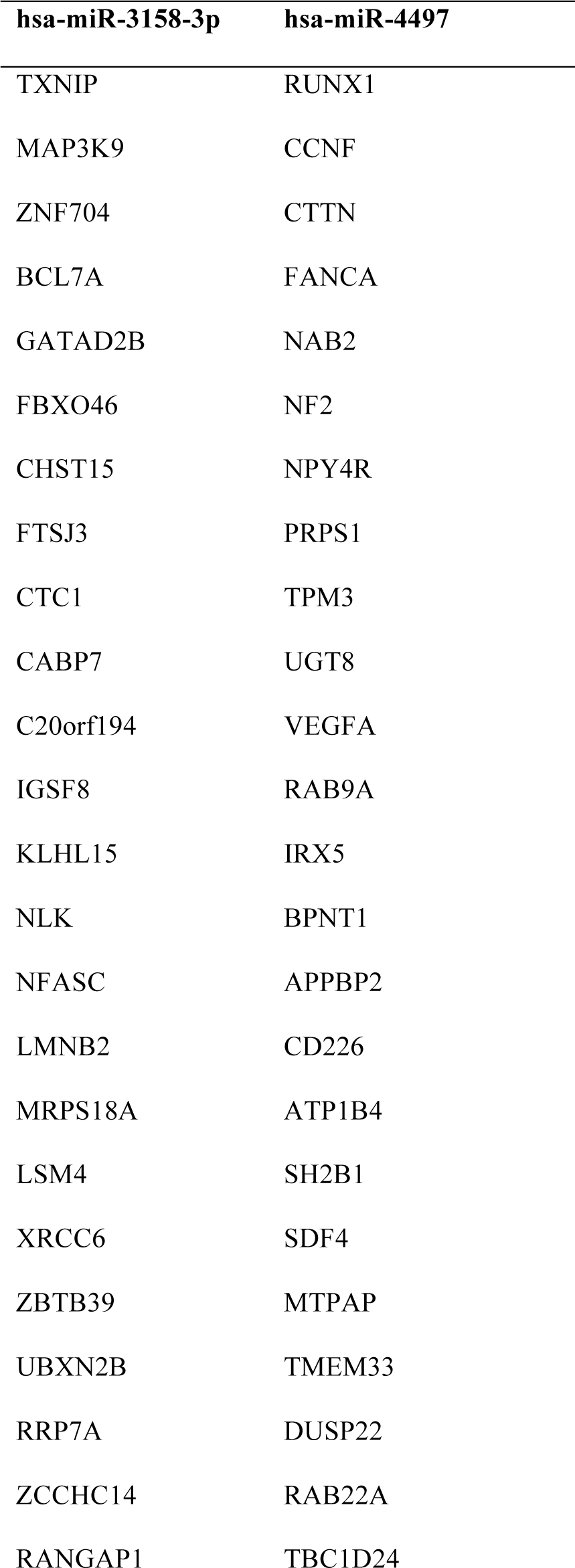
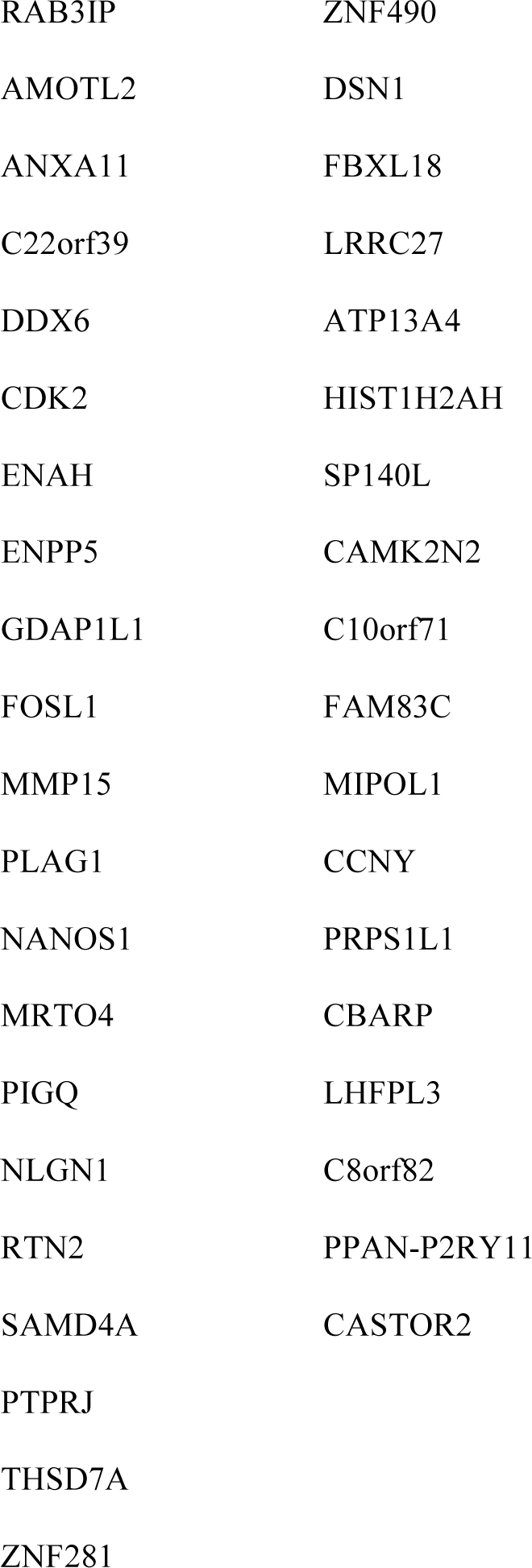
The predicted targets of the two miRNAs (hsa-miR-3158-3p and hsa-miR-4497) validated in children recruited in 2014.

**Table 8:**
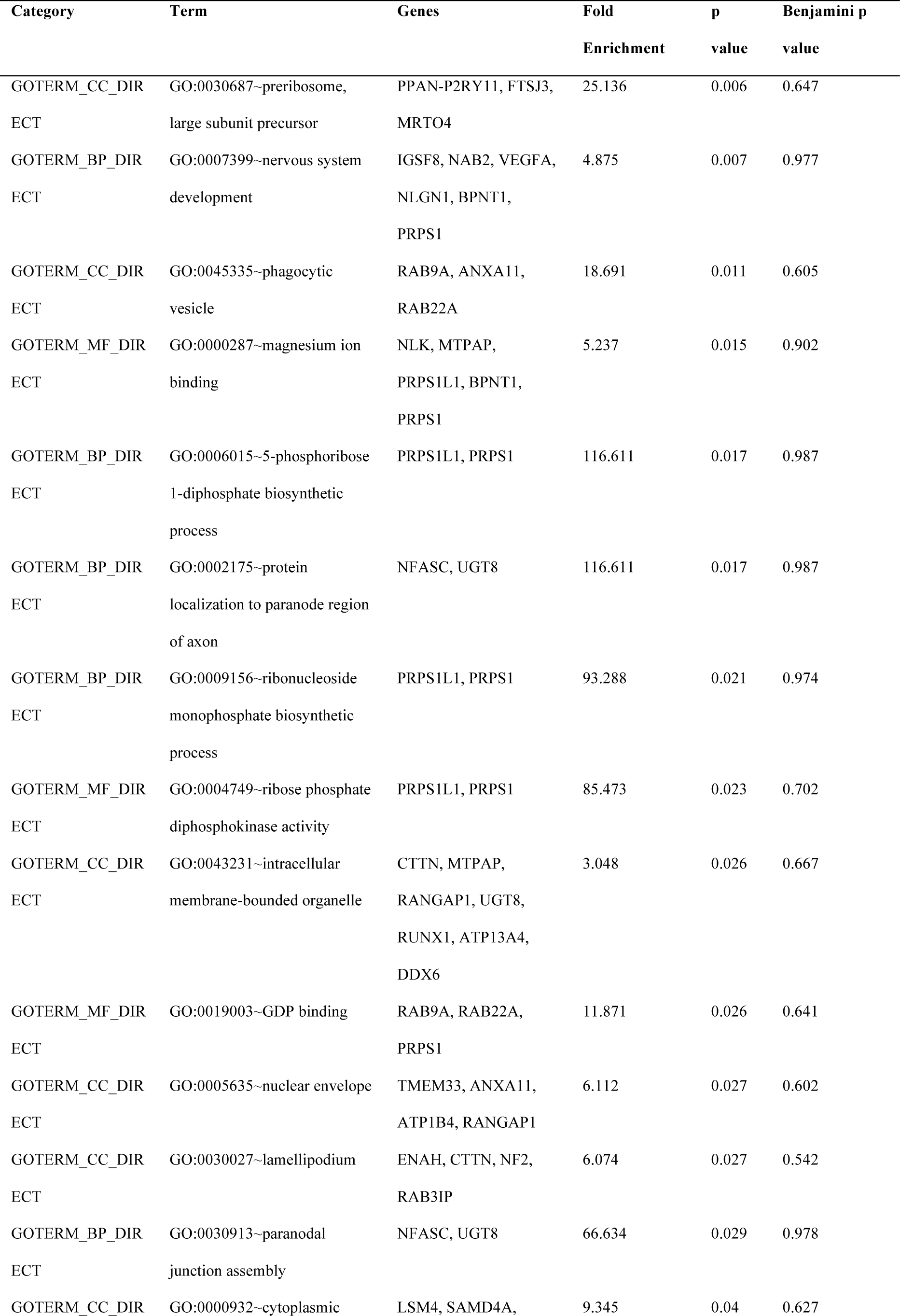
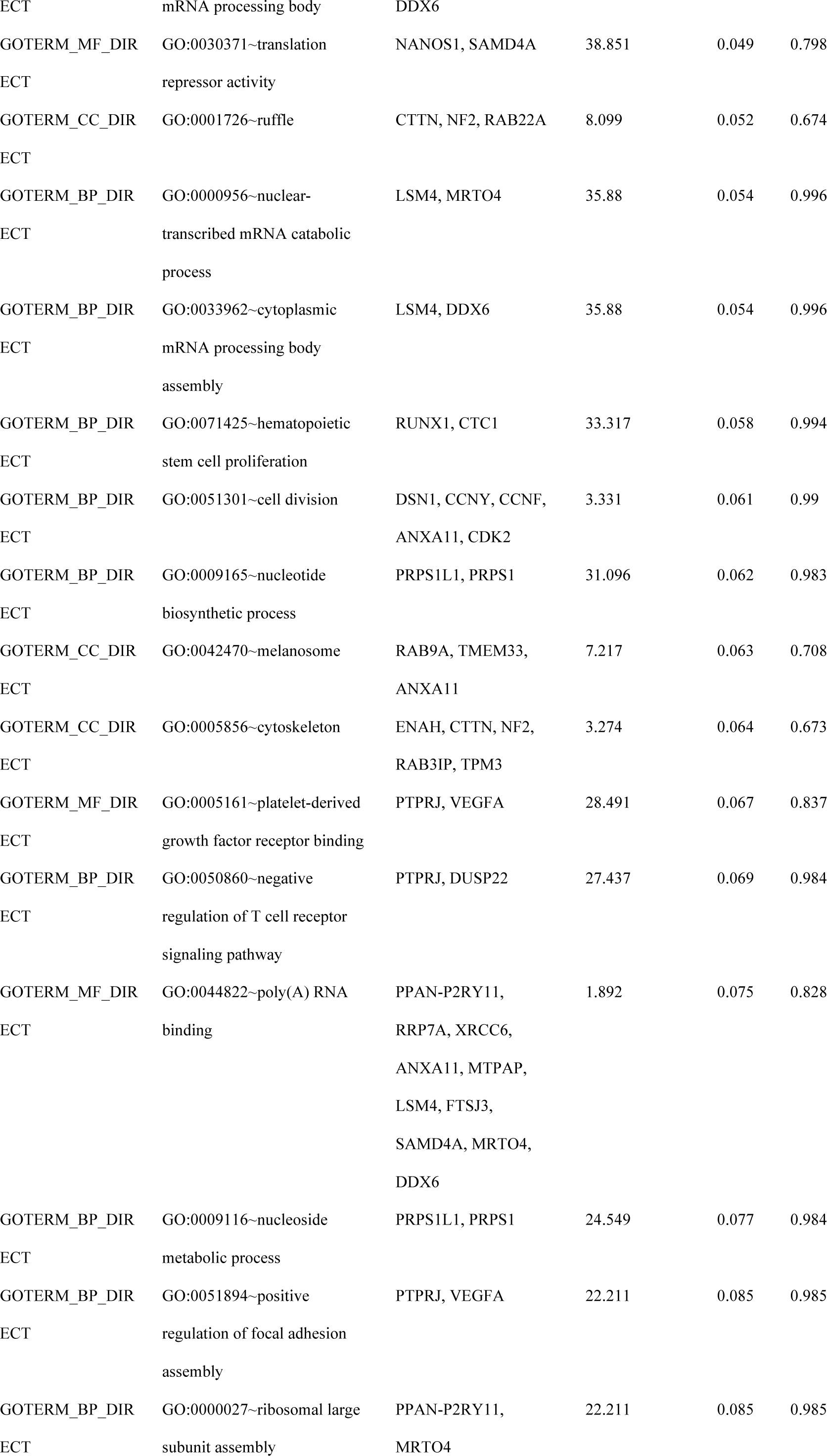
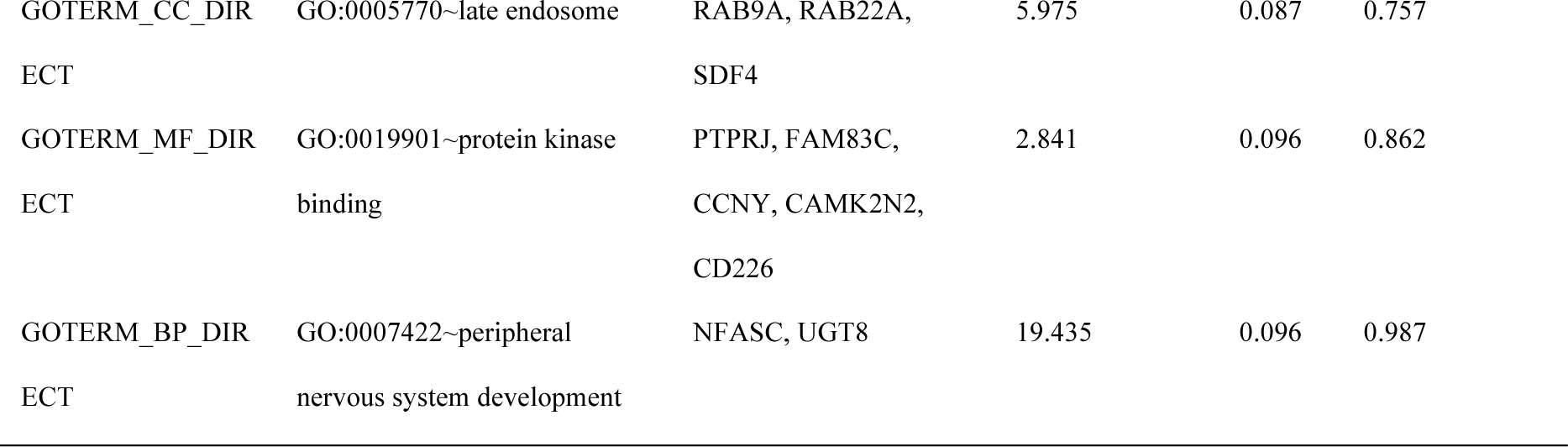
Clustering results of DAVID analysis.

